# Brain-Based Gene Expression of Putative Risk Genes for Anorexia Nervosa in the Human Brain

**DOI:** 10.1101/2022.09.07.22279681

**Authors:** Stuart B. Murray, Jaroslav Rokicki, Alina Sartorius, Adriano Winterton, Ole A. Andreassen, Lars T. Westlye, Jason M. Nagata, Daniel S. Quintana

## Abstract

The etiology of anorexia nervosa (AN) remains elusive. Here, we characterize spatially distributed expression patterns of risk genes for AN in the human brain, developing whole-brain maps of AN gene expression. We found that genes associated with AN are most expressed in the brain, relative to all other body tissue types, and demonstrate gene-specific expression patterns which extend to cerebellar, limbic and basal ganglia structures in particular. fMRI meta-analyses reveal that AN gene expression maps correspond with functional brain activity involved in processing and anticipating appetitive and aversive cues.

---

Anorexia nervosa (AN) is a debilitating and life-threatening psychiatric disorder characterized by self-directed starvation, low weight and emaciation, and an intense fear of weight gain^1^. The pathophysiology of AN, and the biological mechanisms driving the potentially lethal self-directed restriction of food intake, remain elusive. As genome-wide association studies (GWAS) have identified genetic loci associated with complex diseases in humans, a fundamental challenge facing the ‘post-GWAS era’ lies in understanding *how* specific genes confer risk for complex diseases—this is also true of AN. With a stereotypic symptom presentation which remains homogenous across time and culture, elevated risk in first degree relatives of AN probands^2.3^, and substantial twin-based heritability estimates (48-84%)^4-6^, AN appears to be a highly heritable neuropsychiatric phenotype.

Locating the precise source of the genetic risk for AN has historically proven challenging. However, the more recent cumulative aggregation of all available genotyped AN data recently revealed the breakthrough discovery of eight loci reaching genome-wide significance that are associated with nine genes^7^. Elucidating the magnitude and neuroanatomical distribution of the expression of the prioritized genes may offer novel insights into the neurogenetic mechanisms of AN. Specifically, genetic associations at the level of non-disordered human tissue transcriptome may offer important insights into normative gene function, without the confound of clinical epiphenomena common among clinical populations^8^, and postmortem mRNA of human samples in particular has been outlined as the ‘ultimate intermediate phenotype’ to examine neuropsychiatric disorders^9^.

Here we characterize the neuroanatomical distribution of multiarray derived mRNA expression patterns of the nine genes associated with AN^7^ in the non-disordered human brain, and explore the functional relevance of these patterns. To do this, we extracted data from 20,737 protein-coding genes from the Allen Human Brain Atlas (http://human.brain-map.org/), which contains post-mortem brain tissue from six healthy donors (*m* age=42.5 years; *SD*=11.2 years), sampled in 363-946 brain locations within approximately 22 hours of death. First, we assessed the donor-to-donor reproducibility of the nine genes linked to AN in the largest GWAS to date^7^ (**Supplementary Figure 1)** by leveraging the concept of differential stability^10^, which we operationalized as the average Spearman’s correlation between any possible combination of 15 pairs between donors^11^. We found that that five of these genes (*FOXP1, CADM1, CDH10, NCKIPSD, MGMT*) demonstrate strong differential stability, ranking above the 50^th^ percentile of all 20,737 protein-coding genes, indicating reproducible patterning irrespective of sex and ethnicity (**Figure 1**).

**Figure 1:**
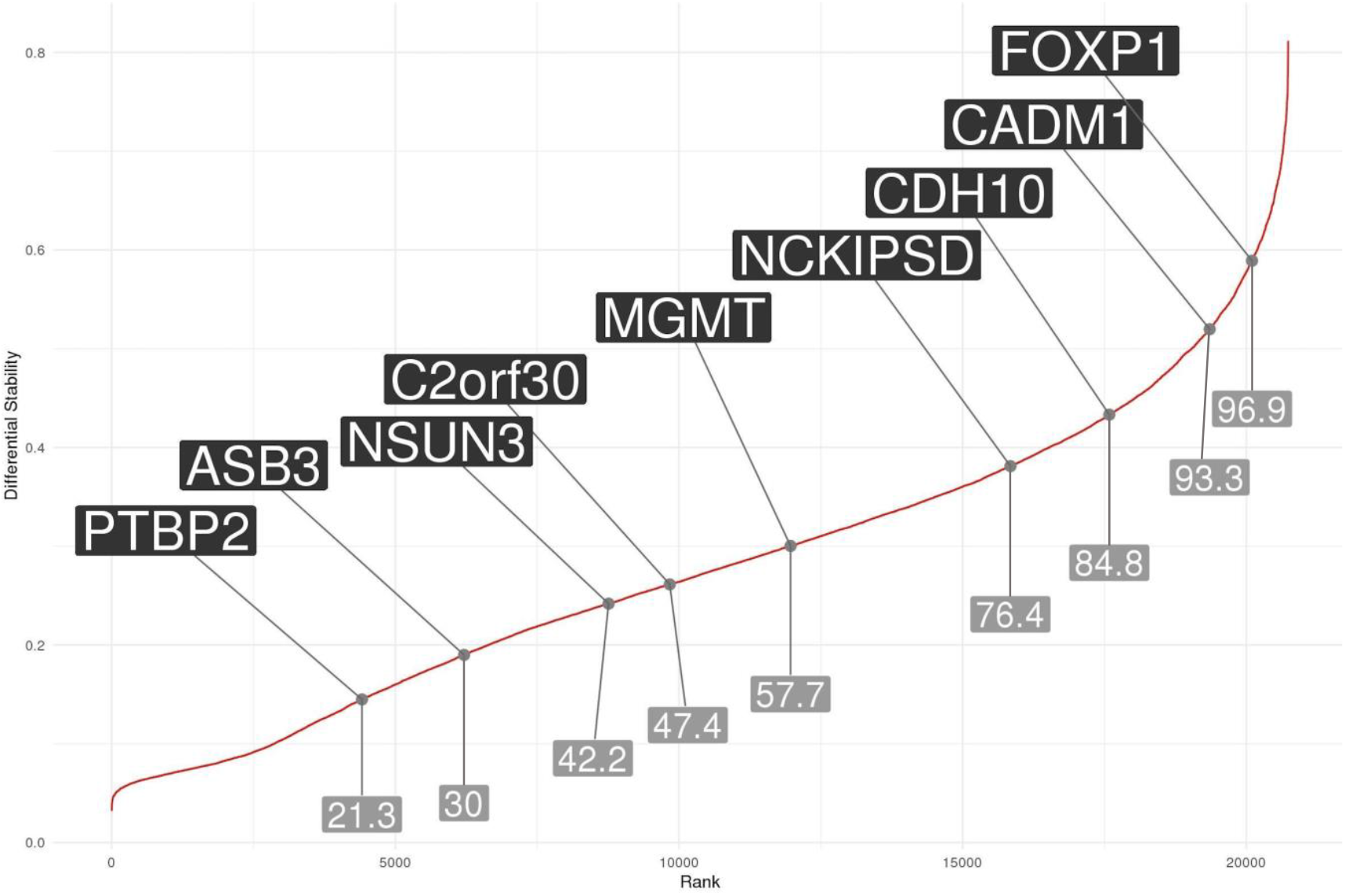
Differential stability of protein coding genes. Differential stability for all protein coding genes (n=20,737) was calculated to assess the similarity of gene expression patterns from donor-to-donor. *FOXP1, CADM1, CDH10, NCKIPSD* and *MGMT* were above the 50^th^ percentile of all genes, suggesting adequate reproducibility.

Next, we created voxel-by-voxel volumetric gene expression maps for the five genes demonstrating strong differential stability and developed a composite brain map representing an average of 6 individuals on the left hemisphere, registered brains to MNI space using ANT’s non-linear registration and averaged so that each gene’s mRNA expression pattern is represented by a single voxel-by-voxel brain map (**Supplementary Figure 2**). One-sample t-tests, corrected for 54 tests using a false discovery rate threshold, were conducted to assess which of the 54 left hemisphere regions from six donor samples expressed mRNA to a significantly greater or lesser degree compared to average mRNA expression across the brain. Of the five genes showing reproducible expression patterns, three demonstrated spatially specific expression patterns which reached statistical significance for specific brain regions (**Figure 2**). Voxel-by-voxel volumetric gene expression maps for the additional six genes are presented in **Supplementary Figure 3**. Importantly, out of sample validation indicates similar expression patterns for all genes of interest in the Genotype-Tissue Expression (GTEx) project database (**Supplementary Figure 4**).

**Figure 2:**
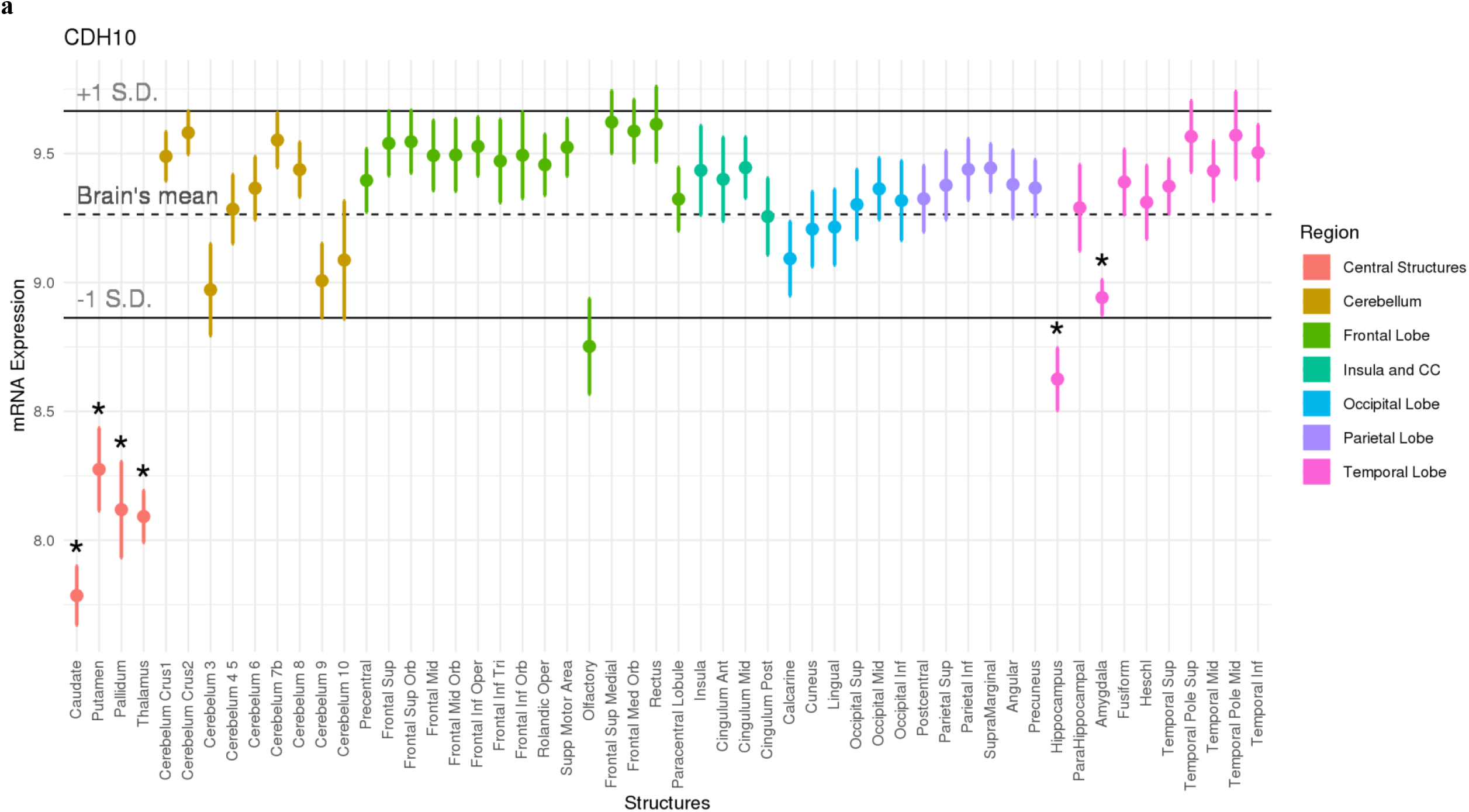
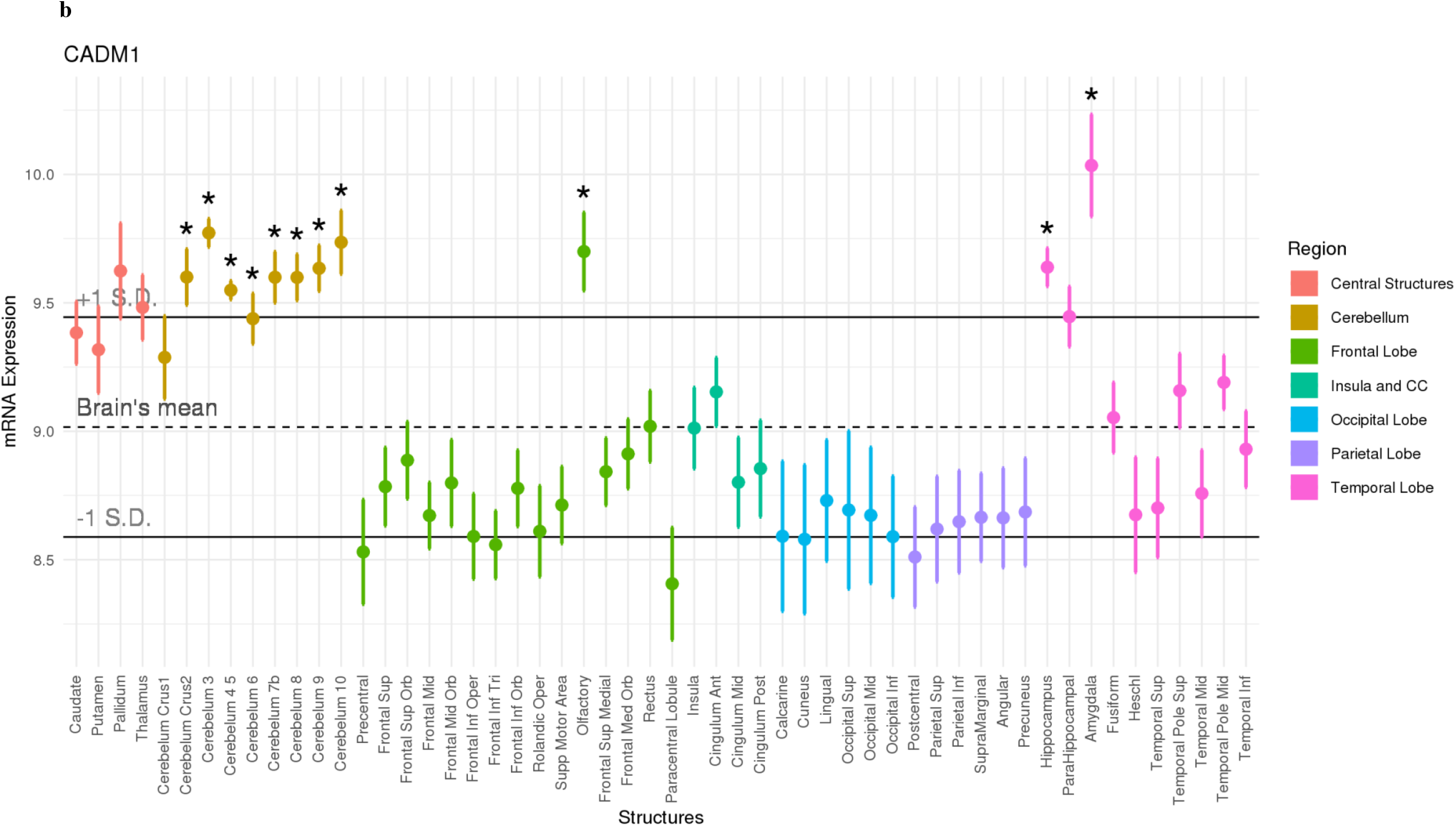
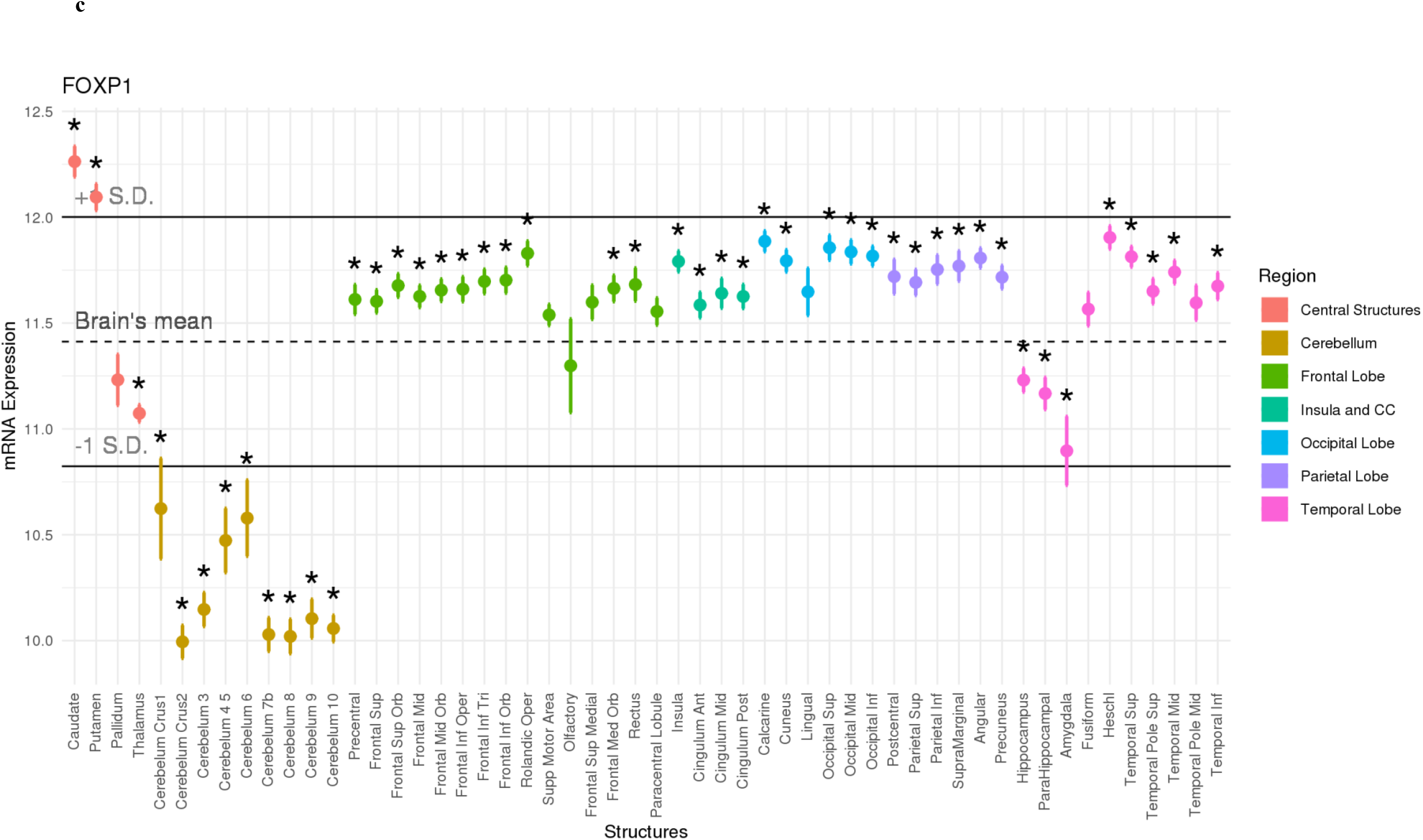
The pathway of gene expression for anorexia nervosa risk genes in the human brain. Each point represents expression from six donors with standard errors for each given brain region for **(a)** CHD10, **(b)** CADM1, and **(c)** FOXP1. Asterisks represent regions of statistically significant over or under expression, relative to the rest of the brain (**\***p<0.05, FDR corrected for 54 tests).

*CADM1* mRNA expression was elevated throughout the cerebellum, the olfactory bulb, and in limbic regions including the hippocampus and amygdala (*p* > 0.05, FDR corrected for 54 tests). Previous GWAS studies have linked *CADM1* to the regulation of body mass and energy homeostasis^12,13^. For instance, risk alleles for obesity in humans are associated with greater mRNA expression of the *CADM1* gene in the cerebellum and hypothalamus^14^, and animal studies have illustrated elevated neuronal expression of *CADM1* in cerebellar, hypothalamic and hippocampal regions in obese mice relative to lean mice^15^. Moreover, the induction of *CADM1* in excitatory neurons facilitates weight gain while paradoxically enhancing energy expenditure^15^. In contrast, the removal of *CADM1* in knockout mice results in a prolonged negative energy balance, rapid weight loss, and prevents weight gain even in the context of extended high fat dietary regimens^15,^ which mirrors the rapid weight loss and profound difficulty reported by those with AN in gaining weight^16,17^. Importantly, the expression of CADM1 in cerebellar and hippocampal regions may be gated by a dynamic interplay between bodyweight and dietary intake, as dietary restriction serves to reduce *CADM1* expression in obese mice, but not in controls^15^.

The *FOXP1* gene, which encodes a transcription factor important for the early development of many organs including the brain^18^, demonstrated highly diffuse patterns of *over*-expression in frontal, occipital, parietal, and temporal regions, and patterns of profound *under*-expression in an array of cerebellar regions, alongside thalamic, hippocampal and amygdalar regions (*p* > 0.05, FDR corrected for 54 tests). In addition, *CDH10* was significantly *under*-expressed in central structures such as the caudate, putamen, pallidum, and thalamus, and temporal regions such as the hippocampus and parahippocampus in the non-disordered human brain (*p* > 0.05, FDR corrected for 54 tests). Interestingly, both *FOXP1* and *CDH10* expression patterns have been linked to the pathophysiology of autism ^19,20^. For instance, overexpression of *FOXP1* has been associated with autism ^20,21^ and related features including language impairment and intellectual disability^22^. Elevated expression of *CDH10* in the frontal cortex has also been associated with autism spectrum disorders^23^. Certainly, AN has been characterized by elevated autistic traits^24,25^ and conceptualized by some as a related endophenotype characterized by cognitive rigidity and social cognition^26,27^. A particular challenge relating to the treatment of AN relates to the high rates of relapse, which is partly underpinned by cognitive and behavioral inflexibility^28^. These data suggest that *FOXP1* and *CDH10* expression may be a plausible mechanism through which cognitive flexibility is altered in those with AN.

Alongside whole-brain gene expression, we additionally assessed gene expression across 30 different body tissue types, by extracting normalized gene expression values (reads per kilo base per million; RPKM) from the GTEx database, via the FUMA platform (**Supplementary Figure 5**). Normalized expression [zero mean of log2(RPKM + 1)] was used to assess differentially expressed gene sets^29^, and Bonferroni adjusted p-values were calculated using two-sided t-tests per gene per tissue against all other tissues. These analyses indicate that the aggregated set of AN risk genes are cumulatively most expressed in the brain, relative to other body tissue types.

To investigate the functional relevance of gene expression patterns, we next investigated how gene-specific gene expression patterns across the whole brain were associated with functional brain activation patterns. We correlated voxel-by-voxel mRNA expression maps for the genes of interest with NeuroSynth (version 0.3.7), performing quantitative reverse inference via large-scale meta-analysis of functional neuroimaging data using mRNA brain expression maps on voxel-by-voxel left hemisphere brain maps, representing the average of the six donors. We assessed the 5 strongest relationships for expression maps for each gene of interest and all available cognitive and mental state maps in the NeuroSynth database.

Expression maps for two of the five differentially stable genes (*CADM1, NCKIPSD*) were highly correlated with functional imaging maps reflecting ‘conditioning’, ‘fear’ and ‘reward’, ranking among the top 0.5% strongest associations for each of these cognitive state activation maps, respectively (**Figure 3**). Moreover, when assessing the relationship between gene expression maps and functional activation maps most associated with specific mental states, these same two genes (*CADM1, NCKIPSD*) were among the 0.5% strongest associations with depression, anxiety, stress and addiction (**Figure 4**). In addition, two of these five genes (*NCKIPSD, MGMT*) were correlated with functional imaging maps reflecting visual processing.

**Figure 3:**
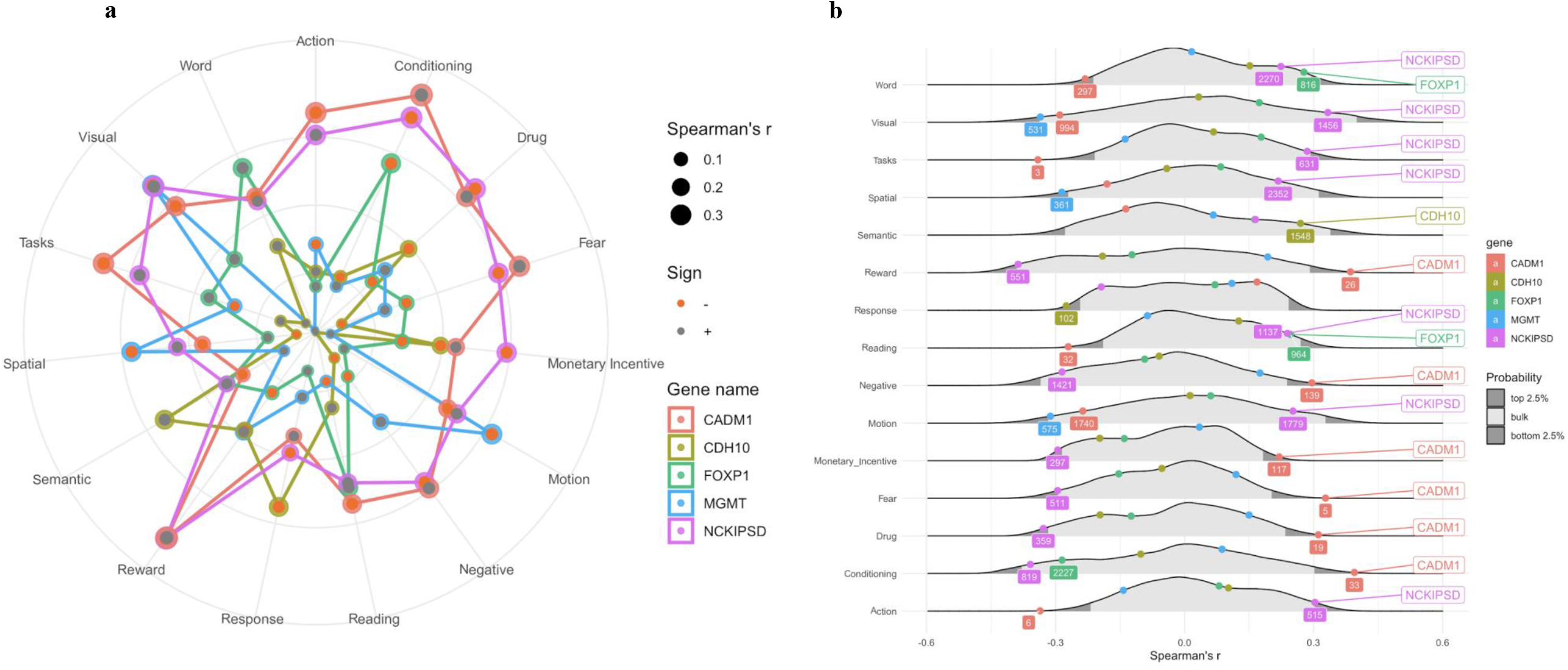
Cognitive state correlates of the five differentially stable genes associated with AN. **a** Cognitive states were meta-analytically decoded from AN risk gene mRNA maps (Supplementary Figure 2) using the NeuroSynth framework. The top five strongest relationships for *CADM1, CDH10, FOXP1, MGMT* and *NCKIPSD* are shown, with duplicates removed. **b** The absolute distribution of Spearman’s correlations between each protein coding gene map (*N*=20,737) and cognitive state maps.

**Figure 4:**
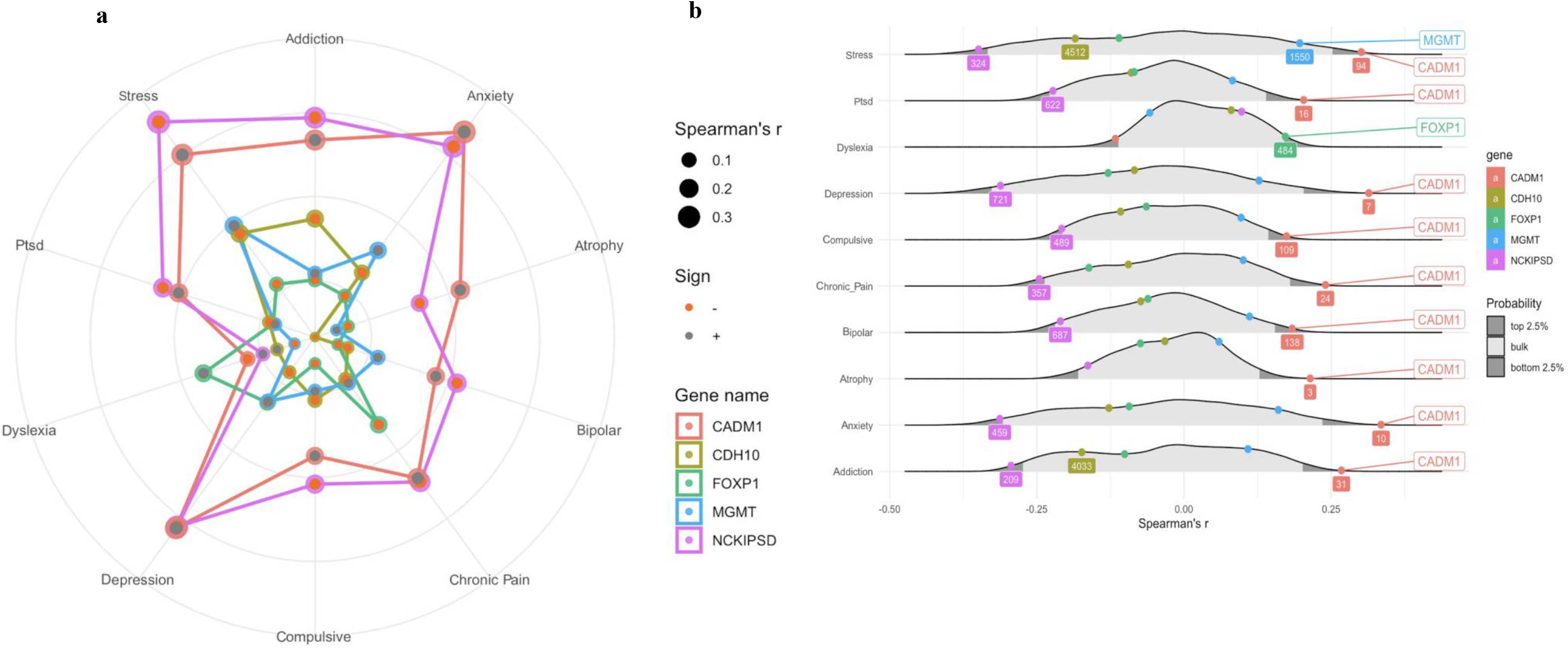
Mental state correlates of the five differentially stable genes associated with AN. **a** Mental states were meta-analytically decoded from AN risk gene mRNA maps (Supplementary Figure 2) using the NeuroSynth framework. The top five strongest relationships for *CADM1, CDH10, FOXP1, MGMT* and *NCKIPSD* are shown, with duplicates removed. **b** The absolute distribution of Spearman’s correlations between each protein coding gene map (*N*=20,737) and mental state maps.

Cumulatively, these results expand recent GWAS findings by offering proof-of-principle demonstration that the genes conferring risk intersect multiple regions and cognitive processes which function abnormally in AN. These findings offer important insights around putative mechanisms through which risk genes associated with AN may confer psychopathology, and how relevant gene expression patterns depend to an extent on both weight and nutritional status which point towards novel targets in elucidating the pathophysiology of AN.

## Methods

### Gene Selection

The largest GWAS of AN to date revealed eight loci which reached genome-wide significance^7^. Extraction of the nearest genes (the nearest gene within the region of linkage disequilibrium ‘friends’ of the lead variant)^7^ resulted in the identification of nine candidate genes.

### Post-Mortem Brain Samples

mRNA distribution data was obtained from the Allen Human Brain Atlas (http://human.brain-map.org/). One donor was a Hispanic female, three donors were Caucasian males, and two donors were African-American males. Mean donor age was 42.5 years (S.D. = 11.2 years), and postmortem brain tissue was collected, on average, 22.3 hours after death. Data collection adhered to ethical guidelines for the collection of human postmortem issue collection, and institutional review board approval was granted at each issue bank and repository that provided tissue samples. Further, informed consent was obtained from each donor’s next-of-kin. For more details regarding data collection procedures, see http://help.brainmap.org/display/humanbrain/Documentation.

### Gene expression data

Gene expression data from 20,737 protein coding genes was collected from the Allen Human Brain Atlas. Each donor brain was sampled in 363-946 locations, either in the left hemisphere only (n = 6), or over both hemispheres (n = 2) using a custom Agilent 8 × 60 K cDNA array chip. Analyses were performed on left hemisphere samples due to a larger sample size. Individual brain maps were non-linearly registered to the MNI152 (Montreal Neurological Institute) template using Advanced Normalization Tools. Next, we extracted region specific statistics for 54 brain regions based on the Automated Anatomical Label (AAL) atlas.

### Donor-to-donor reproducibility of gene expression patterns

Owing to differences in donor sex and ethnicity, we assessed the similarity of gene expression patterns across the six donors by leveraging the concept of differential stability^10^, which is the average Spearman’s correlation between any possible combination of 15 pairs between donors^11^. This method has previously indicated that genes with strong differential stability are highly biologically relevant^10^. For analysis, we selected the probe with the greatest differential stability, which represented the probe with the least amount of spatial variability among donors. The average correlation (Spearman’s r) across 15 pairs of 6 donors’ voxel-by-voxel brain maps were used to calculate differential stability using an approach described by Hawrylycz and colleagues (2015)^10^. Since each voxel-by-voxel map was generated based on a limited and variable number of samples, to calculate the statistical significance of Spearman’s coefficients we calculated p-values between two donors based on the smallest number of samples out of the two. That is, if one donor had samples from 353 locations and another one from 456, we would use the 353 samples to calculate a p-value for the pair.

### Out-of-sample validation

The Genotype-Tissue Expression (GTEx) project database was used for independent sample validation. Although this dataset provides gene expression data from fewer brain regions (i.e., 10) compared to the Allen dataset, the data is derived from a larger dataset of donors (mean sample size for mRNA expression across brain regions = 131.7, range = 88–173). Median gene expression profiles from 10 distinct brain regions were extracted for the above-specified 20 genes of interest from the GTEx database and median values were calculated for these same 10 regions from the Allen dataset. For independent sample validation, the rank-order correlation of gene expression between the Allen and GTEx datasets using the 10 brain regions reported in the GTEx database was calculated.

### Voxel-by-voxel gene expression maps

To create novel voxel-by-voxel volumetric expression maps, we first marked all the sample locations and expression values in native image space^11,30^. To interpolate missing voxels, we labeled brain borders with the sample expression value that had the closest distance to a given border point (Supplementary Fig. 2). Next, we divided the space between scattered points into simplices based on Delaunay triangulation, then linearly interpolated each simplex with values to yield a completed map. All maps were computed in Matlab 2014a (The Mathworks Inc., Natick, MA, USA). We then created a composite brain map representing an average of 6 individuals on the left hemisphere, registered brains to MNI space using ANT’s non-linear registration and averaged so that each gene’s mRNA is represented by a single voxel-by-voxel brain map.

### Cognitive state correlates

The Neurosynth framework has collated neuroimaging data from over 14,000 fMRI studies (database version 0.7, released July, 2018). While this framework can be used to develop meta-analytic brain activation maps for specific cognitive states (i.e., stress, learning, reward) using forward inference, it may also be leveraged to “decode” cognitive states on a given activation map, via reverse inference^11^. We correlated voxel-by-voxel mRNA expression maps for the genes of interest with NeuroSynth (version 0.3.7), performing quantitative reverse inference via large-scale meta-analysis of functional neuroimaging data using mRNA brain expression maps on voxel-by-voxel left hemisphere brain maps, representing the average of the six donors. Next, we modified the NeuroSynth package to calculate Spearman’s correlation coefficient instead of the default Pearson’s correlation coefficient. To test the specificity of these cognitive states, we extracted association Z maps, which reflect Z-scores of the association between the presence of activation and the presence of a cognitive feature. We assessed the 5 strongest relationships for expression maps for each gene of interest and all available cognitive state maps in the Neurosynth database. Additionally, we calculated Spearman’s correlation between all 20,737 genes and association Z score maps and ranked them from largest to smallest.

### Statistical analysis of gene expression data

The R statistical package (version 3.3.2) was used for statistical analysis. One-sample t-tests (two-tailed) were conducted to assess which of the 54 left hemisphere regions from six donor samples expressed mRNA to a significantly greater or lesser degree compared to average mRNA expression across the brain. To correct for multiple tests (54 in total), reported p-values were adjusted using a false discovery rate (FDR) threshold. Cohen’s d values for one-sample t-tests were calculated to yield a measure of effect size.

To assess gene expression across various body tissues, normalized gene expression values (reads per kilo base per million; RPKM) were extracted from the GTEx database, via the FUMA platform. As described by Watanabe and colleagues^29^, normalized expression [zero mean of log2(RPKM + 1)] was used to assess differentially expressed gene sets. Bonferroni adjusted p-values are then calculated using two-sided t-tests per gene per tissue against all other tissues. Genes with a Bonferroni adjusted p-value < 0.05 and absolute log fold change ≥0.58 were categorized as a differentially expressed gene set in a given tissue type. The presented -log10 p-values represent results from hypergeometric tests, which were used to assess if genes of interest were overrepresented in differentially expressed gene sets in specific tissues.

Similarly, hypergeometric tests were used to assess if genes of interest were overrepresented in gene sets reported in the GWAS catalog and gene sets associated with behavioral and cognitive state processes reported within GO biological processes gene sets within Molecular Signatures Database, using 20,119 protein coding genes as the background set. *p*-values were Benjamini-Hochberg adjusted for all genesets reported in the GO biological processes dataset and GWAS catalog, respectively.

## Data Availability

All data produced in the present study are available upon reasonable request to the authors

