## Supplementary material for "Brain-Based Gene Expression of Putative Risk Genes for Anorexia Nervosa in the Human Brain": Online Supplement

**Supplementary Figure 1**: The differential stability of protein-coding genes ($n$=20,737) was calculated to assess the gene expression patterns from donor-to-donor. Each individual association for NSUN3, NCKIPSD, CDH10, FOXP1, CADM1 was statistically significant, suggesting stable donor-to-donor gene expression patterns. All but one individual association was significant for ERLEC1, whereas PTBP2 and ASB3 were characterized by reduced differential stability across donors. * $p$>0.001.

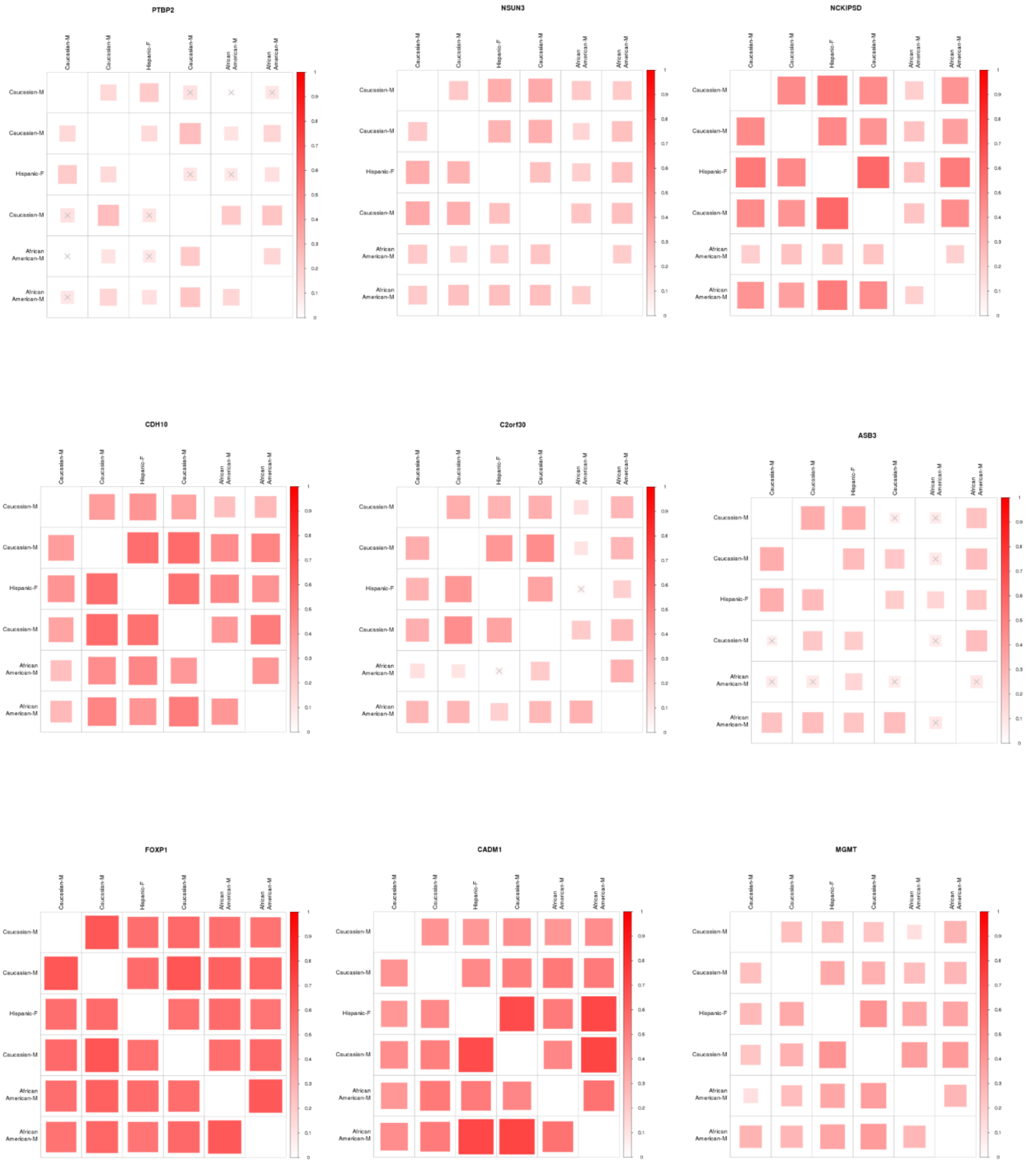

**Supplementary Figure 2**: Voxel-by-voxel brain gene expression maps for the nine risk genes for AN. mRNA expression maps were created using left hemisphere data from the Allen Human Brain Atlas dataset.

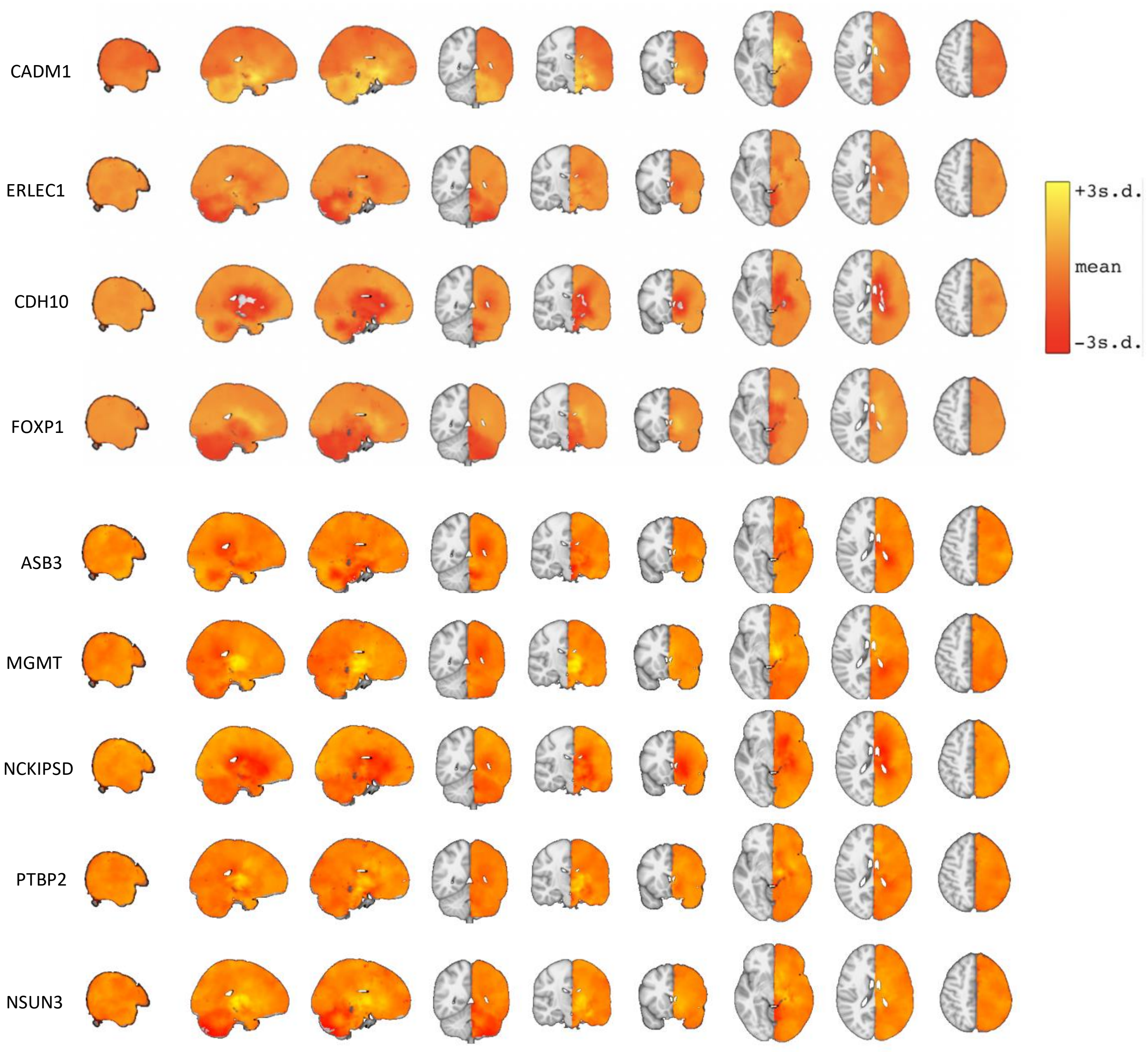

**Supplementary Figure 3**: The pathway of gene expression for anorexia nervosa risk genes in the human brain. Each point represents expression from six donors with standard errors for each given brain region for **(a)** NCKIPSD, **(b)** MGMT, and **(c)** C2orf30, **(d)** NSUN3, **(e)**ASB3, and **(f)** PTBP2. Asterisks represent regions of statistically significant over or under expression, relative to the rest of the brain (*p<0.05, FDR corrected for 54 tests).

**a**

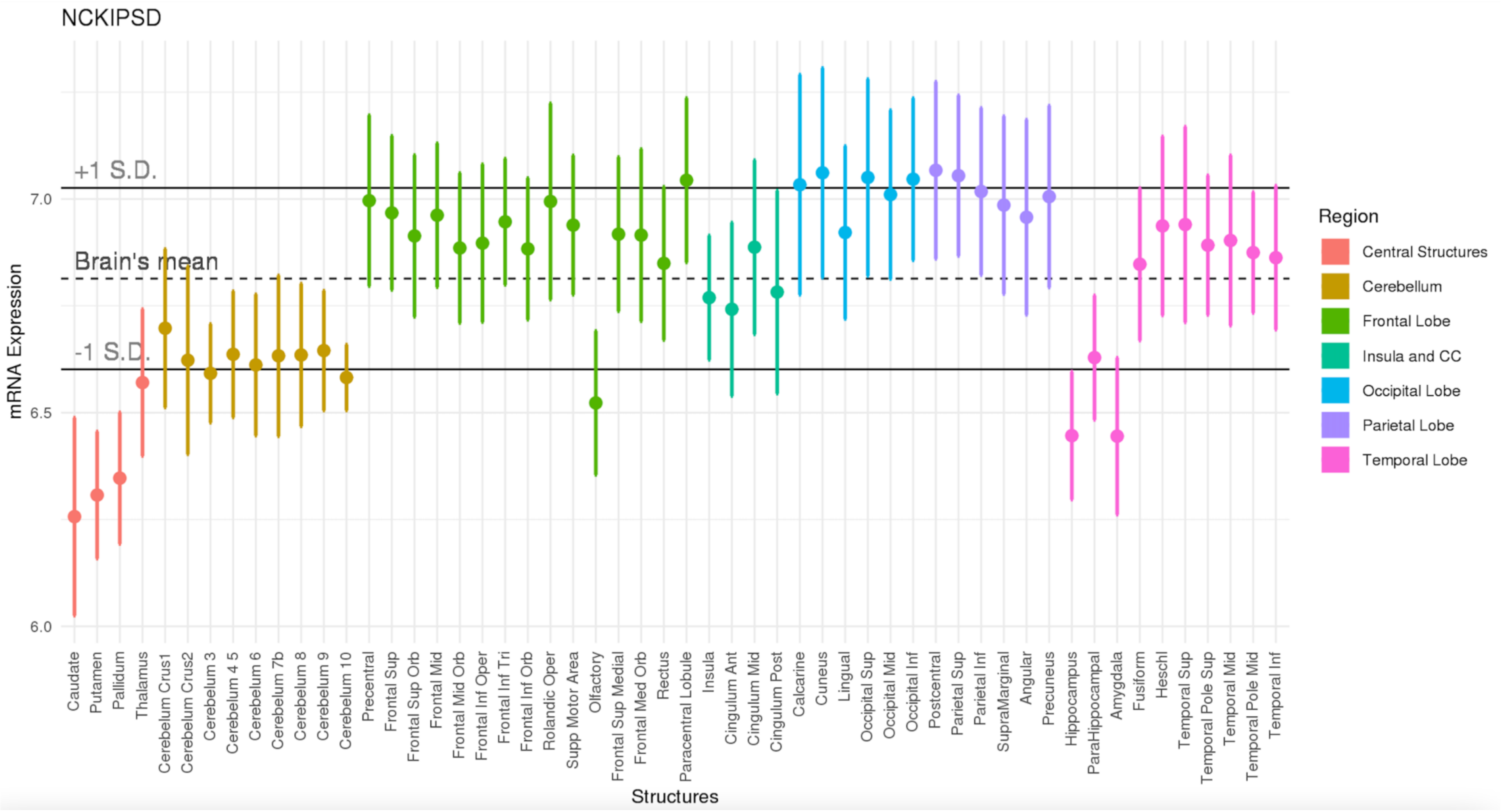

**b**

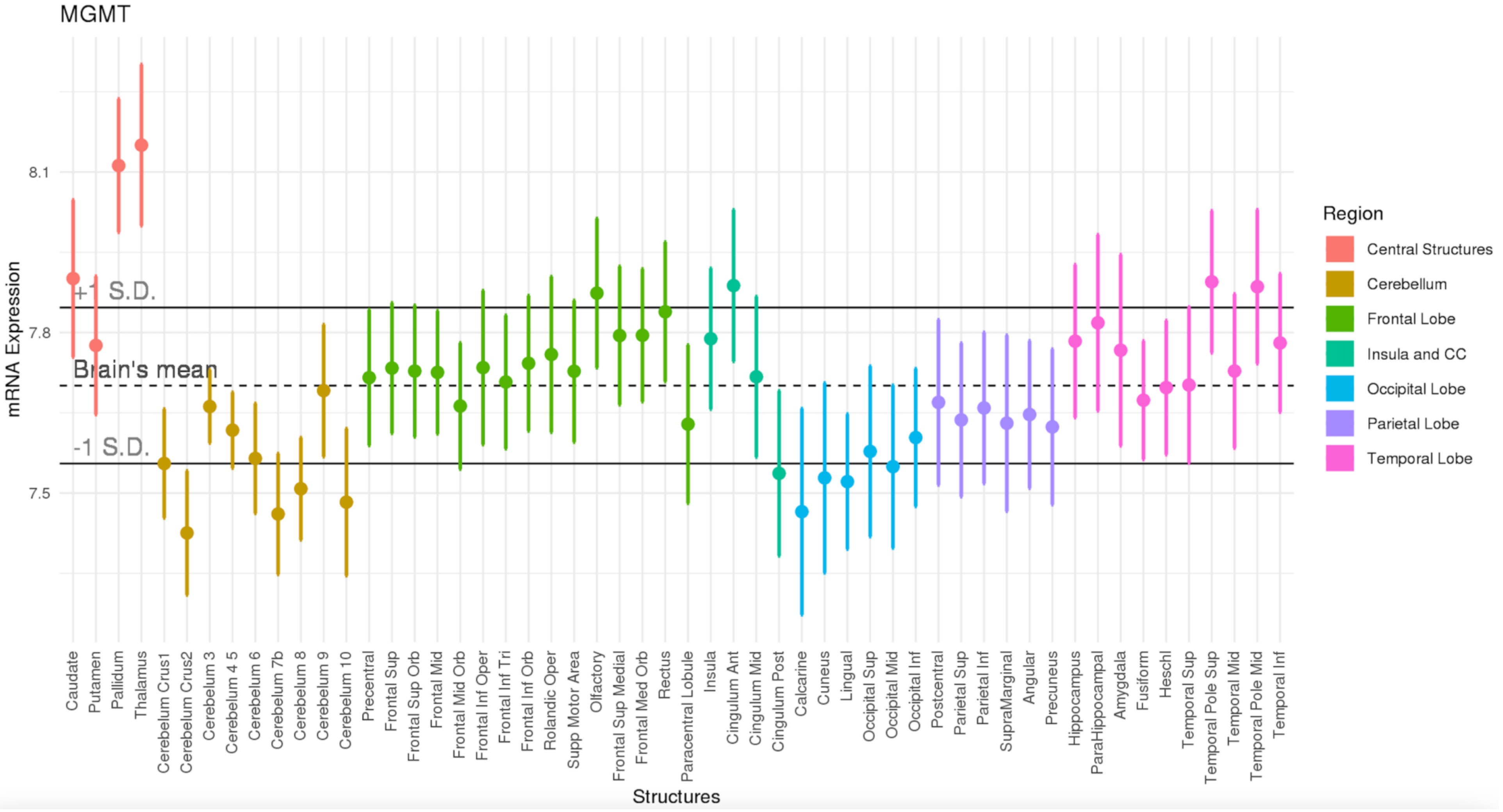

MGMT
mRNA Expression
8.1
7.8
7.5
+1 S.D.
Brain's mean
-1 S.D.
Structures
Caudate
Putamen
Pallidum
Thalamus
Cerebelum Crus1
Cerebelum Crus2
Cerebelum 3
Cerebelum 4 5
Cerebelum 6
Cerebelum 7b
Cerebelum 8
Cerebelum 9
Cerebelum 10
Precentral
Frontal Sup
Frontal Sup Orb
Frontal Mid
Frontal Mid Orb
Frontal Inf Oper
Frontal Inf Tri
Frontal Inf Orb
Rolandic Oper
Supp Motor Area
Olfactory
Frontal Sup Medial
Frontal Med Orb
Rectus
Paracentral Lobule
Insula
Cingulum Ant
Cingulum Mid
Cingulum Post
Calcarine
Cuneus
Lingual
Occipital Sup
Occipital Mid
Occipital Inf
Postcentral
Parietal Sup
Parietal Inf
SupraMarginal
Angular
Precuneus
Hippocampus
ParaHippocampal
Amygdala
Fusiform
Heschl
Temporal Sup
Temporal Pole Sup
Temporal Mid
Temporal Pole Mid
Temporal Inf
Region
Central Structures
Cerebellum
Frontal Lobe
Insula and CC
Occipital Lobe
Parietal Lobe
Temporal Lobe

**c**

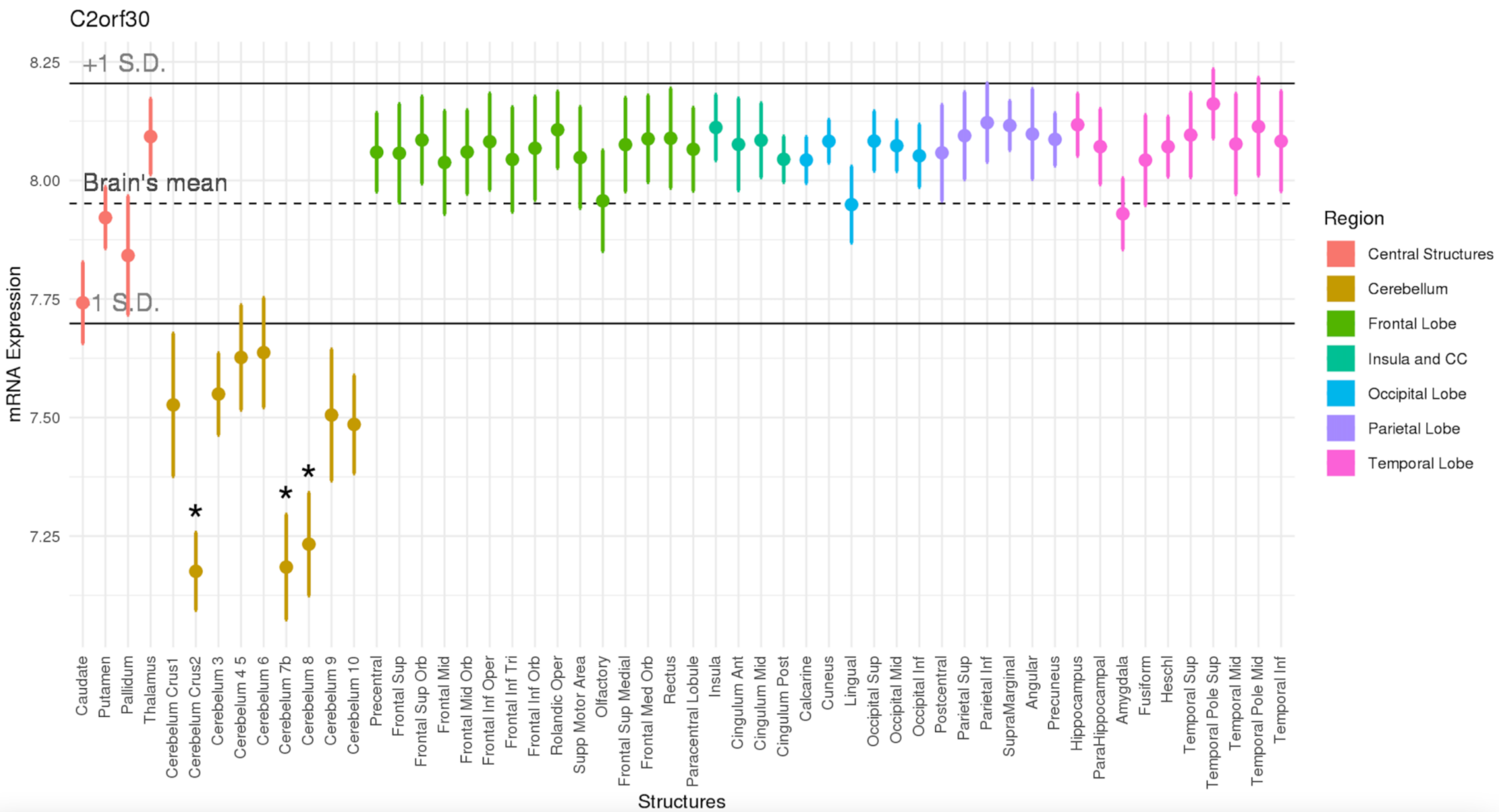
C2orf30
8.25
8.00
7.75
7.50
7.25
+1 S.D.
Brain's mean
-1 S.D.
mRNA Expression
*
*
*
Caudate
Putamen
Pallidum
Thalamus
Cerebelum Crus1
Cerebelum Crus2
Cerebelum 3
Cerebelum 4 5
Cerebelum 6
Cerebelum 7b
Cerebelum 8
Cerebelum 9
Cerebelum 10
Precentral
Frontal Sup
Frontal Sup Orb
Frontal Mid
Frontal Mid Orb
Frontal Inf Oper
Frontal Inf Tri
Frontal Inf Orb
Rolandic Oper
Supp Motor Area
Olfactory
Frontal Sup Medial
Frontal Med Orb
Rectus
Paracentral Lobule
Insula
Cingulum Ant
Cingulum Mid
Cingulum Post
Calcarine
Cuneus
Lingual
Occipital Sup
Occipital Mid
Occipital Inf
Postcentral
Parietal Sup
Parietal Inf
SupraMarginal
Angular
Precuneus
Hippocampus
ParaHippocampal
Amygdala
Fusiform
Heschl
Temporal Sup
Temporal Pole Sup
Temporal Mid
Temporal Pole Mid
Temporal Inf
Structures
Region
Central Structures
Cerebellum
Frontal Lobe
Insula and CC
Occipital Lobe
Parietal Lobe
Temporal Lobe

**d**

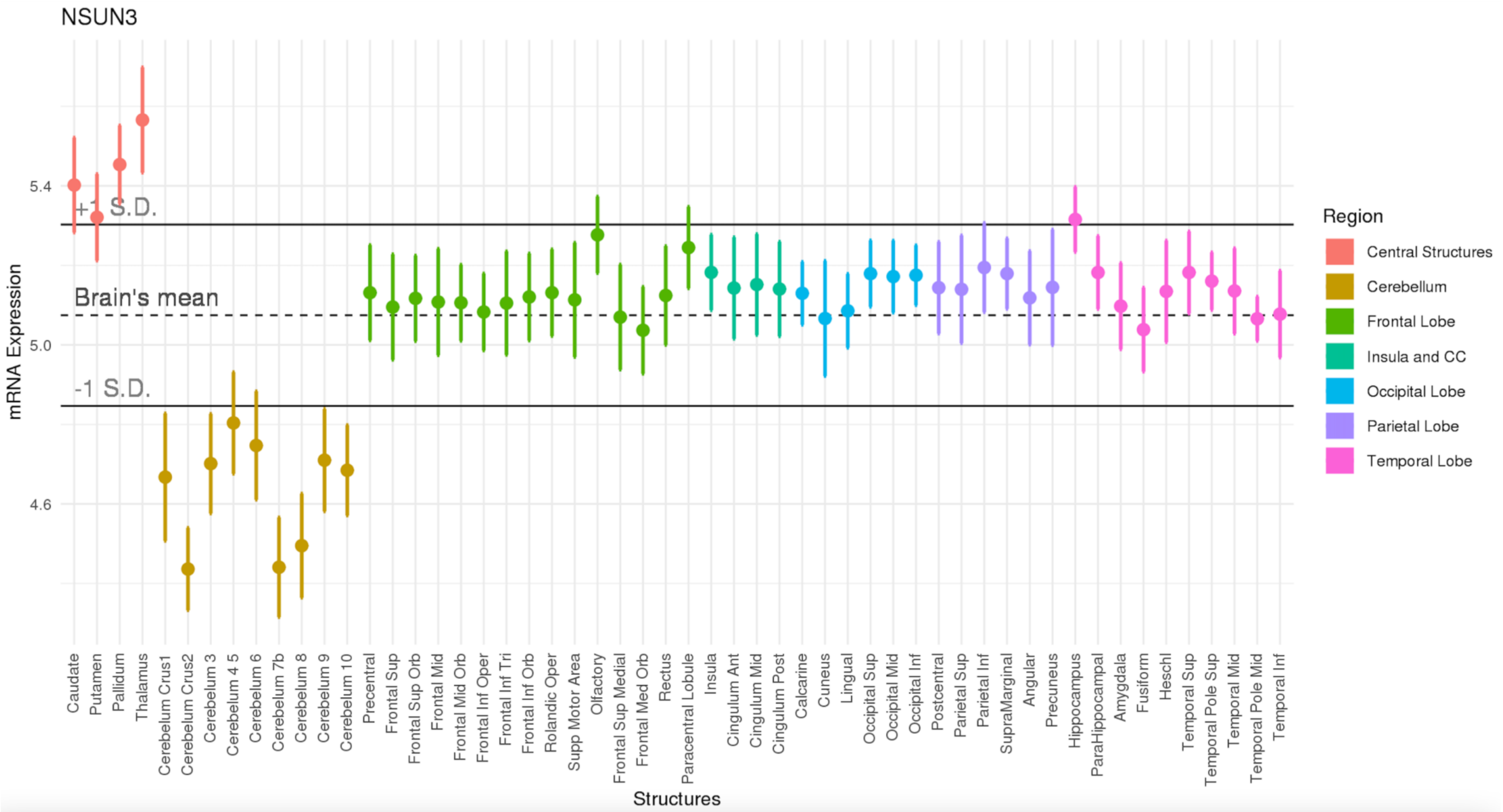
NSUN3
mRNA Expression
5.4
5.0
4.6
+1 S.D.
Brain's mean
-1 S.D.
Structures
Caudate
Putamen
Pallidum
Thalamus
Cerebelum Crus1
Cerebelum Crus2
Cerebelum 3
Cerebelum 4 5
Cerebelum 6
Cerebelum 7b
Cerebelum 8
Cerebelum 9
Cerebelum 10
Precentral
Frontal Sup
Frontal Sup Orb
Frontal Mid
Frontal Mid Orb
Frontal Inf Oper
Frontal Inf Tri
Frontal Inf Orb
Rolandic Oper
Supp Motor Area
Olfactory
Frontal Sup Medial
Frontal Med Orb
Rectus
Paracentral Lobule
Insula
Cingulum Ant
Cingulum Mid
Cingulum Post
Calcarine
Cuneus
Lingual
Occipital Sup
Occipital Mid
Occipital Inf
Postcentral
Parietal Sup
Parietal Inf
SupraMarginal
Angular
Precuneus
Hippocampus
ParaHippocampal
Amygdala
Fusiform
Heschl
Temporal Sup
Temporal Pole Sup
Temporal Mid
Temporal Pole Mid
Temporal Inf
Region
Central Structures
Cerebellum
Frontal Lobe
Insula and CC
Occipital Lobe
Parietal Lobe
Temporal Lobe

e

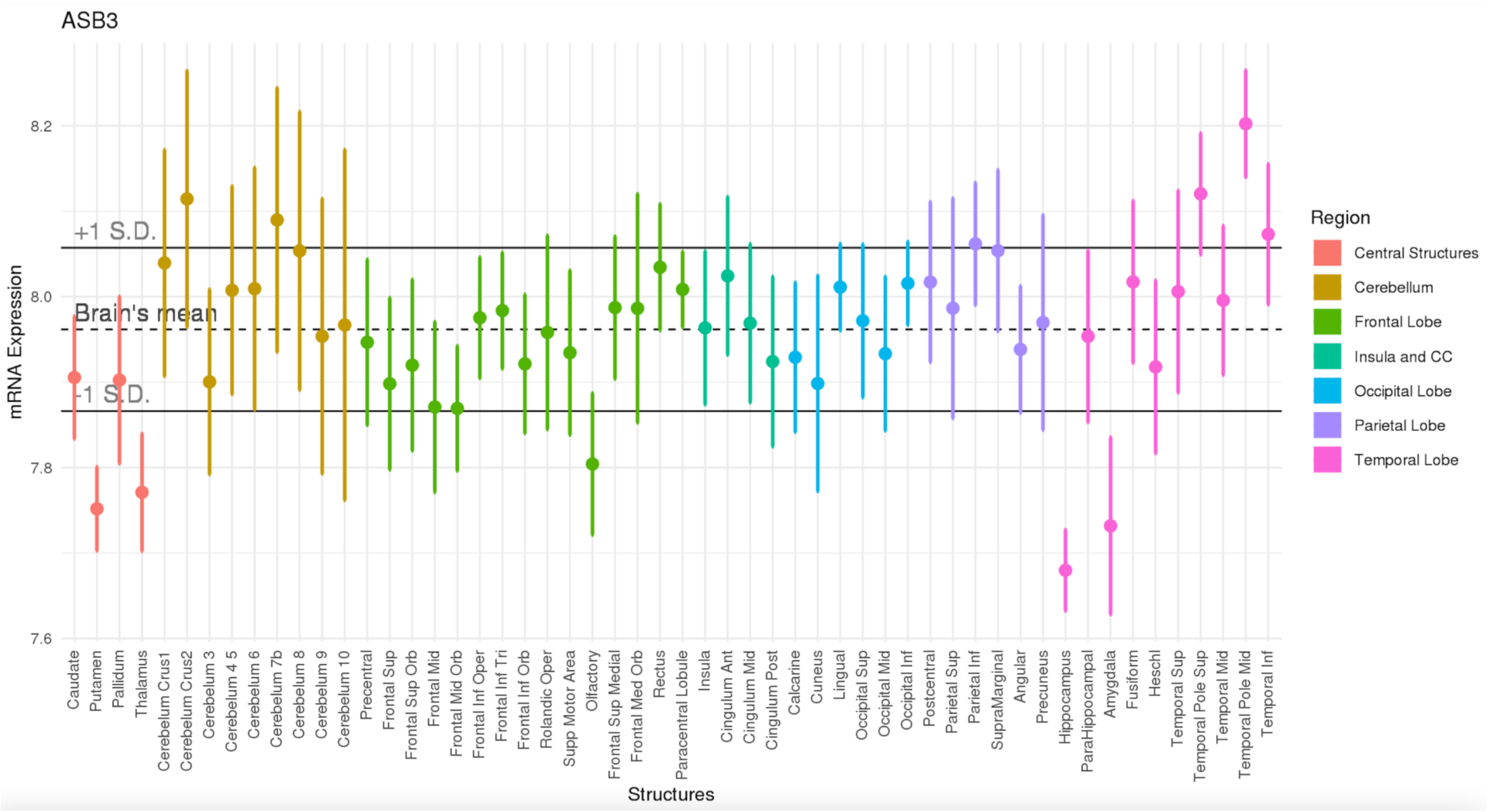
ASB3
mRNA Expression
Structures
+1 S.D.
Brain's mean
-1 S.D.
8.2
8.0
7.8
7.6
Caudate
Putamen
Pallidum
Thalamus
Cerebelum Crus1
Cerebelum Crus2
Cerebelum 3
Cerebelum 4 5
Cerebelum 6
Cerebelum 7b
Cerebelum 8
Cerebelum 9
Cerebelum 10
Precentral
Frontal Sup
Frontal Sup Orb
Frontal Mid
Frontal Mid Orb
Frontal Inf Oper
Frontal Inf Tri
Frontal Inf Orb
Rolandic Oper
Supp Motor Area
Olfactory
Frontal Sup Medial
Frontal Med Orb
Rectus
Paracentral Lobule
Insula
Cingulum Ant
Cingulum Mid
Cingulum Post
Calcarine
Cuneus
Lingual
Occipital Sup
Occipital Mid
Occipital Inf
Postcentral
Parietal Sup
Parietal Inf
SupraMarginal
Angular
Precuneus
Hippocampus
ParaHippocampal
Amygdala
Fusiform
Heschl
Temporal Sup
Temporal Pole Sup
Temporal Mid
Temporal Pole Mid
Temporal Inf
Region
Central Structures
Cerebellum
Frontal Lobe
Insula and CC
Occipital Lobe
Parietal Lobe
Temporal Lobe

f

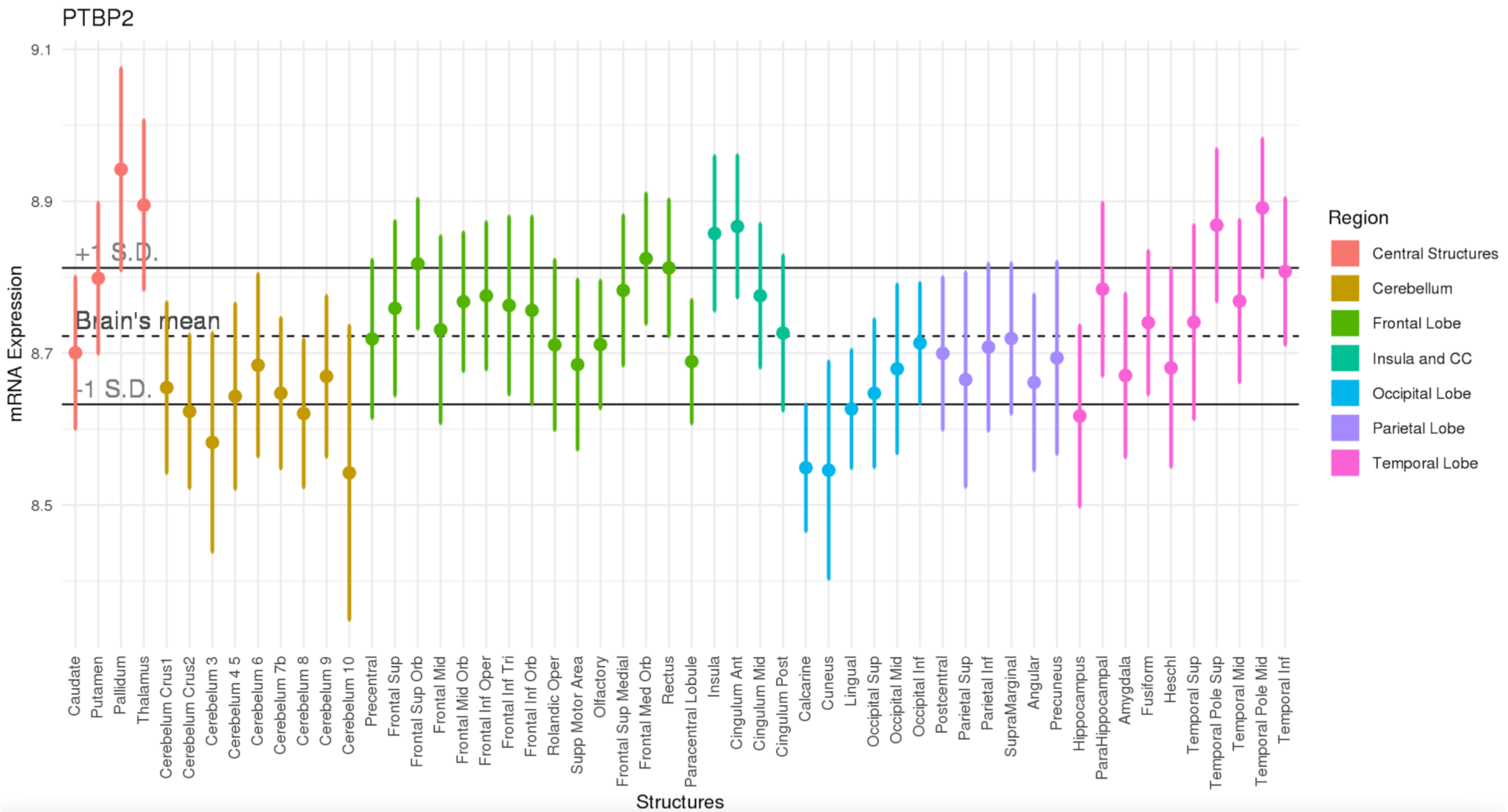

PTBP2
9.1
8.9
8.7
8.5
+1 S.D.
Brain's mean
-1 S.D.
mRNA Expression
Caudate
Putamen
Pallidum
Thalamus
Cerebelum Crus1
Cerebelum Crus2
Cerebelum 3
Cerebelum 4 5
Cerebelum 6
Cerebelum 7b
Cerebelum 8
Cerebelum 9
Cerebelum 10
Precentral
Frontal Sup
Frontal Sup Orb
Frontal Mid
Frontal Mid Orb
Frontal Inf Oper
Frontal Inf Tri
Frontal Inf Orb
Rolandic Oper
Supp Motor Area
Olfactory
Frontal Sup Medial
Frontal Med Orb
Rectus
Paracentral Lobule
Insula
Cingulum Ant
Cingulum Mid
Cingulum Post
Calcarine
Cuneus
Lingual
Occipital Sup
Occipital Mid
Occipital Inf
Postcentral
Parietal Sup
Parietal Inf
SupraMarginal
Angular
Precuneus
Hippocampus
ParaHippocampal
Amygdala
Fusiform
Heschl
Temporal Sup
Temporal Pole Sup
Temporal Mid
Temporal Pole Mid
Temporal Inf
Structures
Region
Central Structures
Cerebellum
Frontal Lobe
Insula and CC
Occipital Lobe
Parietal Lobe
Temporal Lobe

**Supplementary Figure 5:** Out of sample replication of volumetric gene expression maps for the nine genes linked to anorexia nervosa in the human brain. Each point represents expression in six brain regions from the Genotype-Tissue Expression (GTEx) project database, for **(a)** FOXP1, **(b)** CADM1, **(c)** CDH10, **(d)** NCKIPSD, **(e)** MGMT, **(f)** C2orf30, **(g)** NSUN3, **(h)**ASB3, and **(i)** PTBP2. Asterisks represent regions of statistically significant over or under expression, relative to the rest of the brain (***p<0.05, FDR corrected for 10 tests).

**a**

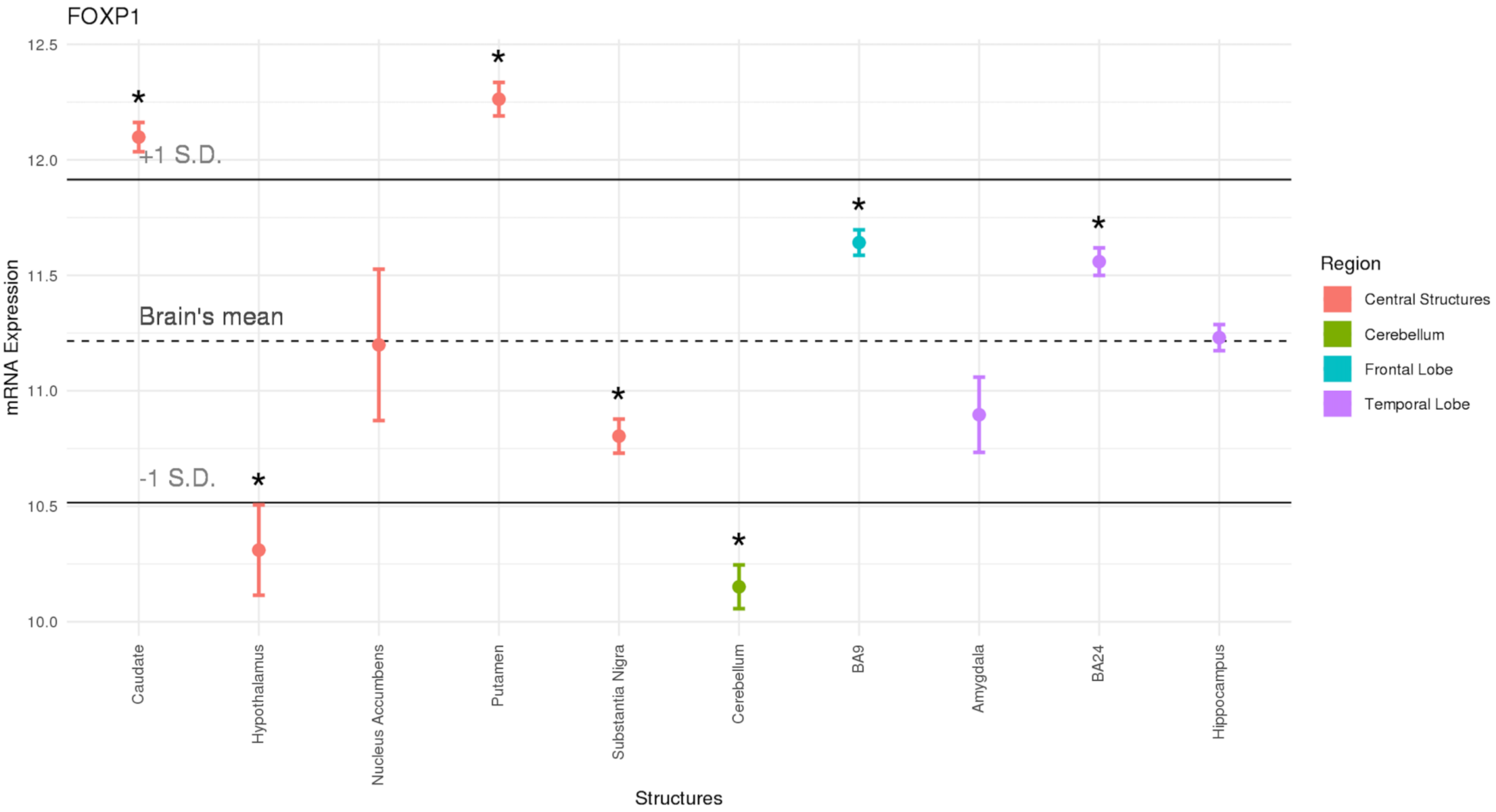

**b**

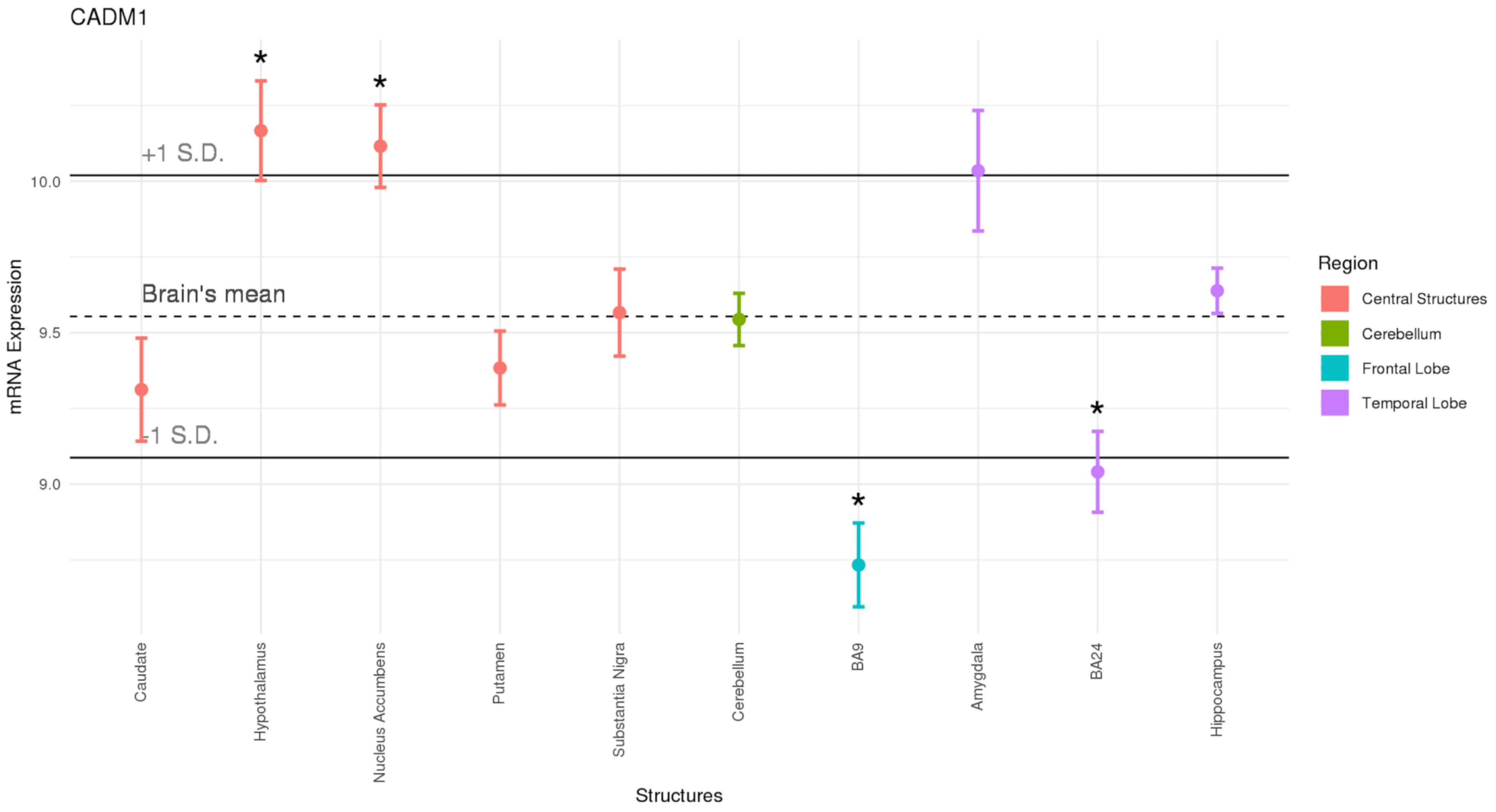
CADM1
+1 S.D.
Brain's mean
-1 S.D.
10.0
9.5
9.0
mRNA Expression
Caudate
Hypothalamus
Nucleus Accumbens
Putamen
Substantia Nigra
Cerebellum
BA9
Amygdala
BA24
Hippocampus
Structures
Region
Central Structures
Cerebellum
Frontal Lobe
Temporal Lobe

c

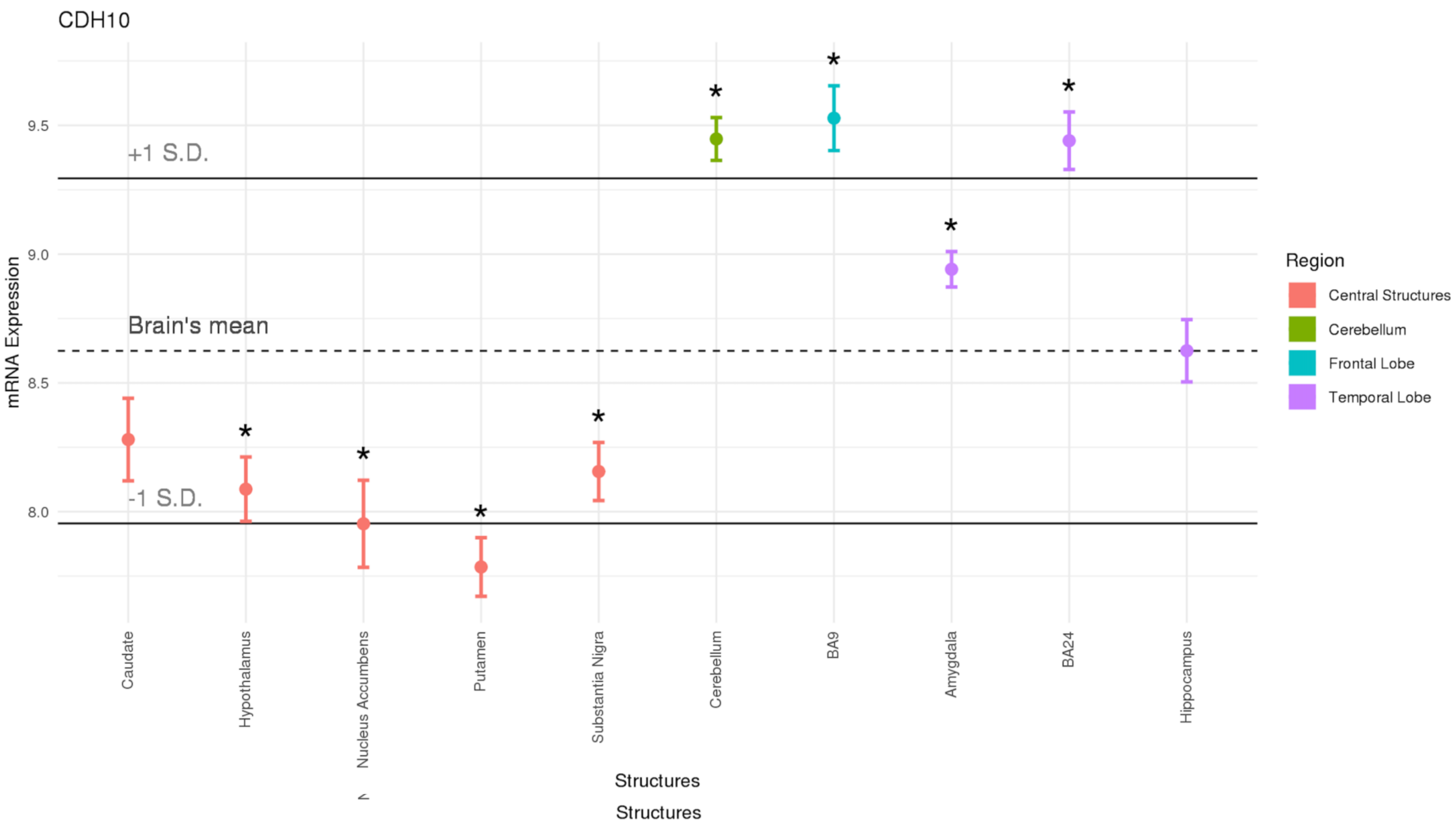
CDH10
mRNA Expression
9.5
9.0
8.5
8.0
+1 S.D.
Brain's mean
-1 S.D.
Caudate
Hypothalamus
Nucleus Accumbens
Putamen
Substantia Nigra
Cerebellum
BA9
Amygdala
BA24
Hippocampus
Structures
Structures
Region
Central Structures
Cerebellum
Frontal Lobe
Temporal Lobe

d

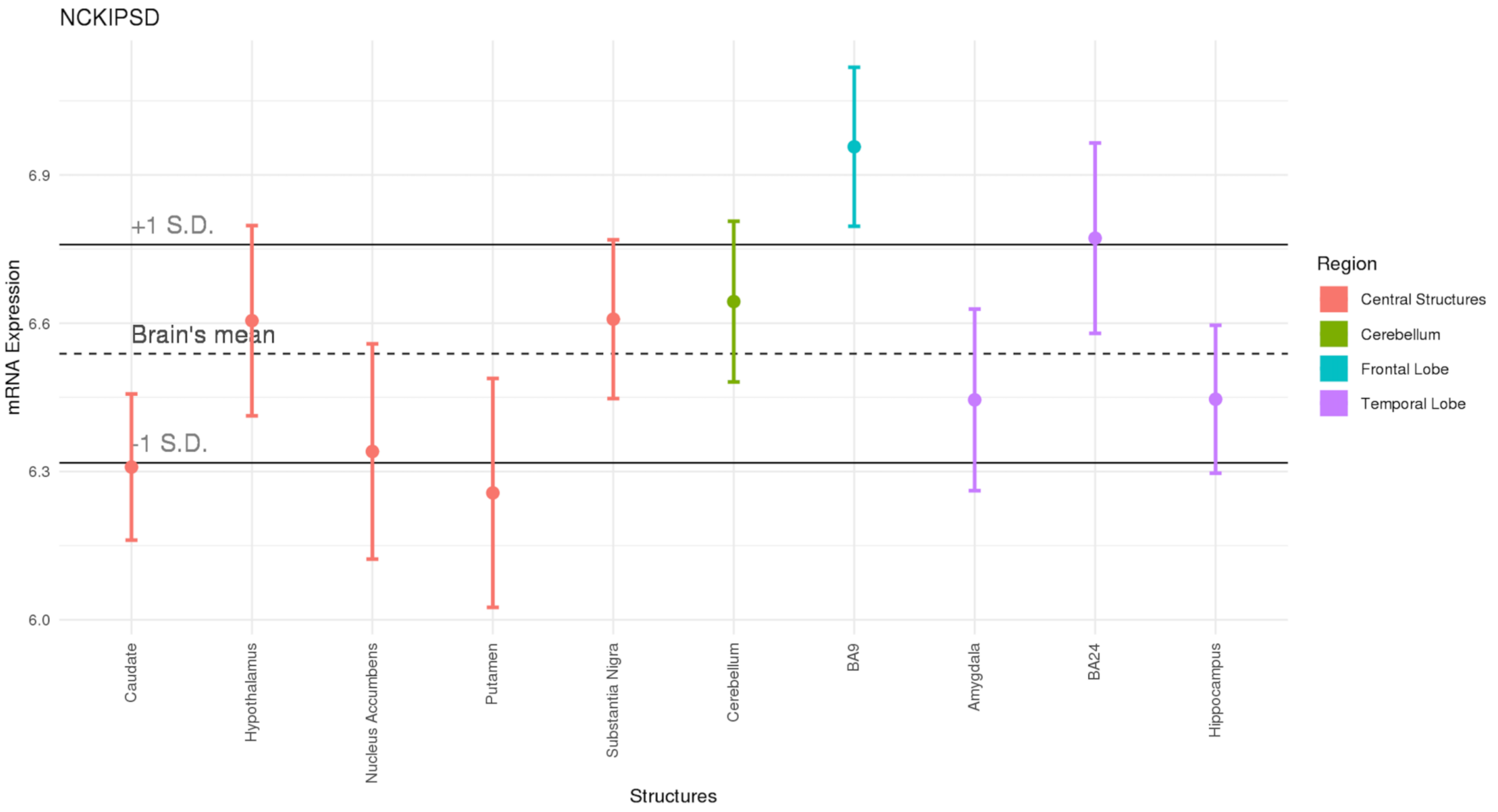
NCKIPSD
+1 S.D.
Brain's mean
-1 S.D.
mRNA Expression
6.9
6.6
6.3
6.0
Caudate
Hypothalamus
Nucleus Accumbens
Putamen
Substantia Nigra
Cerebellum
BA9
Amygdala
BA24
Hippocampus
Structures
Region
Central Structures
Cerebellum
Frontal Lobe
Temporal Lobe

e

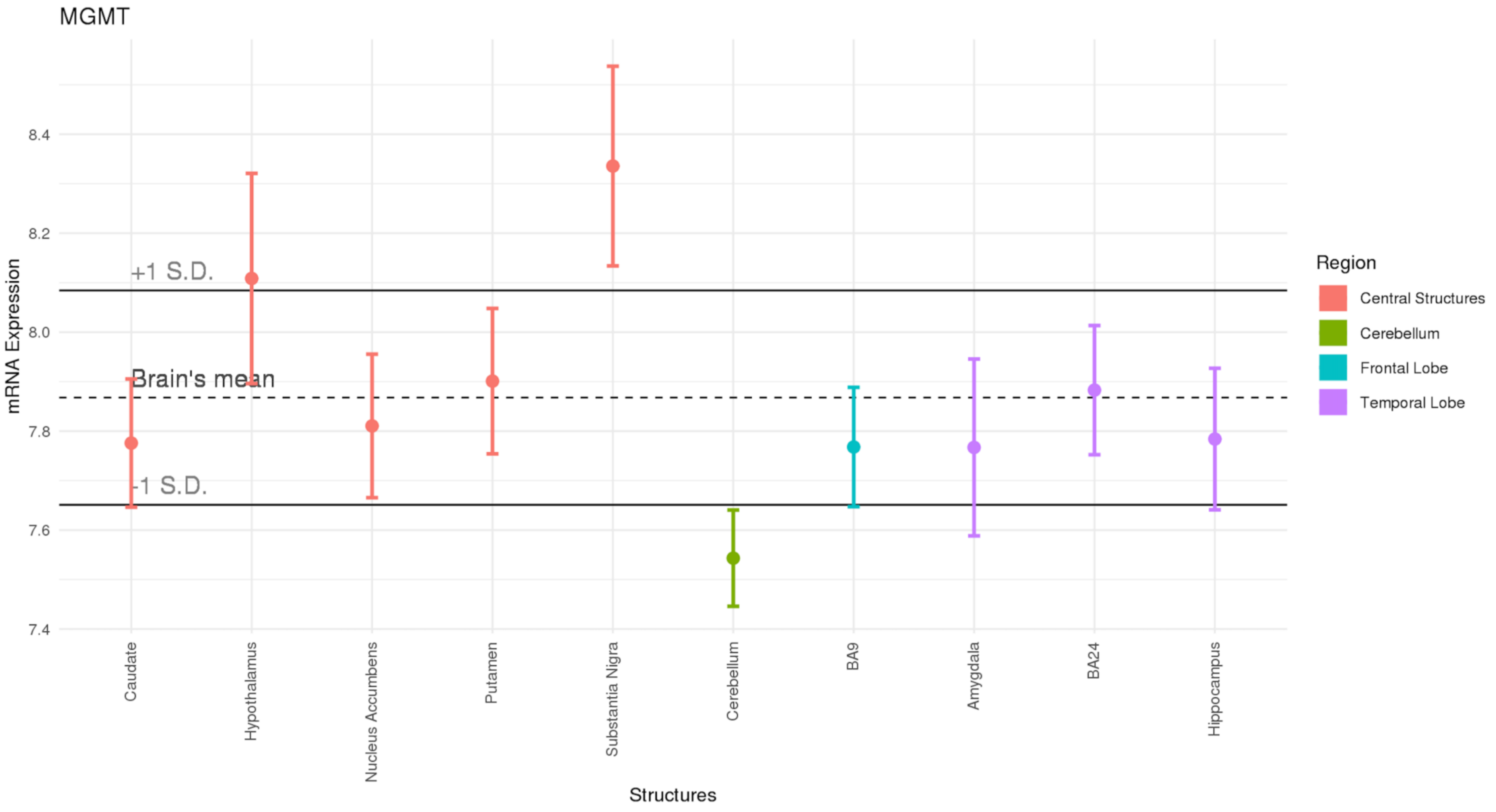
MGMT
mRNA Expression
8.4
8.2
8.0
7.8
7.6
7.4
+1 S.D.
Brain's mean
-1 S.D.
Caudate
Hypothalamus
Nucleus Accumbens
Putamen
Substantia Nigra
Cerebellum
BA9
Amygdala
BA24
Hippocampus
Structures
Region
Central Structures
Cerebellum
Frontal Lobe
Temporal Lobe

f

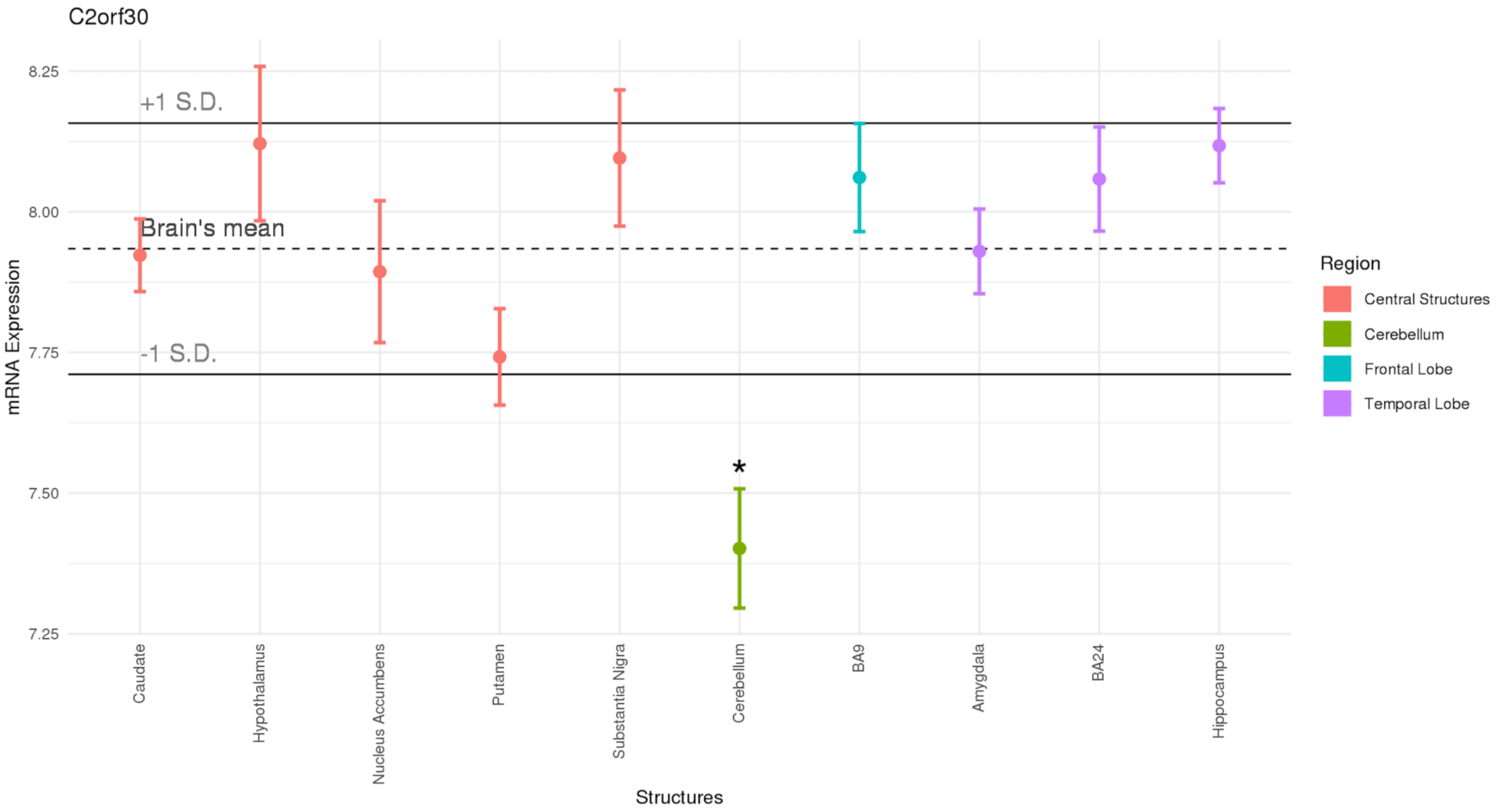
C2orf30
mRNA Expression
8.25
8.00
7.75
7.50
7.25
+1 S.D.
Brain's mean
-1 S.D.
*
Caudate
Hypothalamus
Nucleus Accumbens
Putamen
Substantia Nigra
Cerebellum
BA9
Amygdala
BA24
Hippocampus
Structures
Region
Central Structures
Cerebellum
Frontal Lobe
Temporal Lobe

g

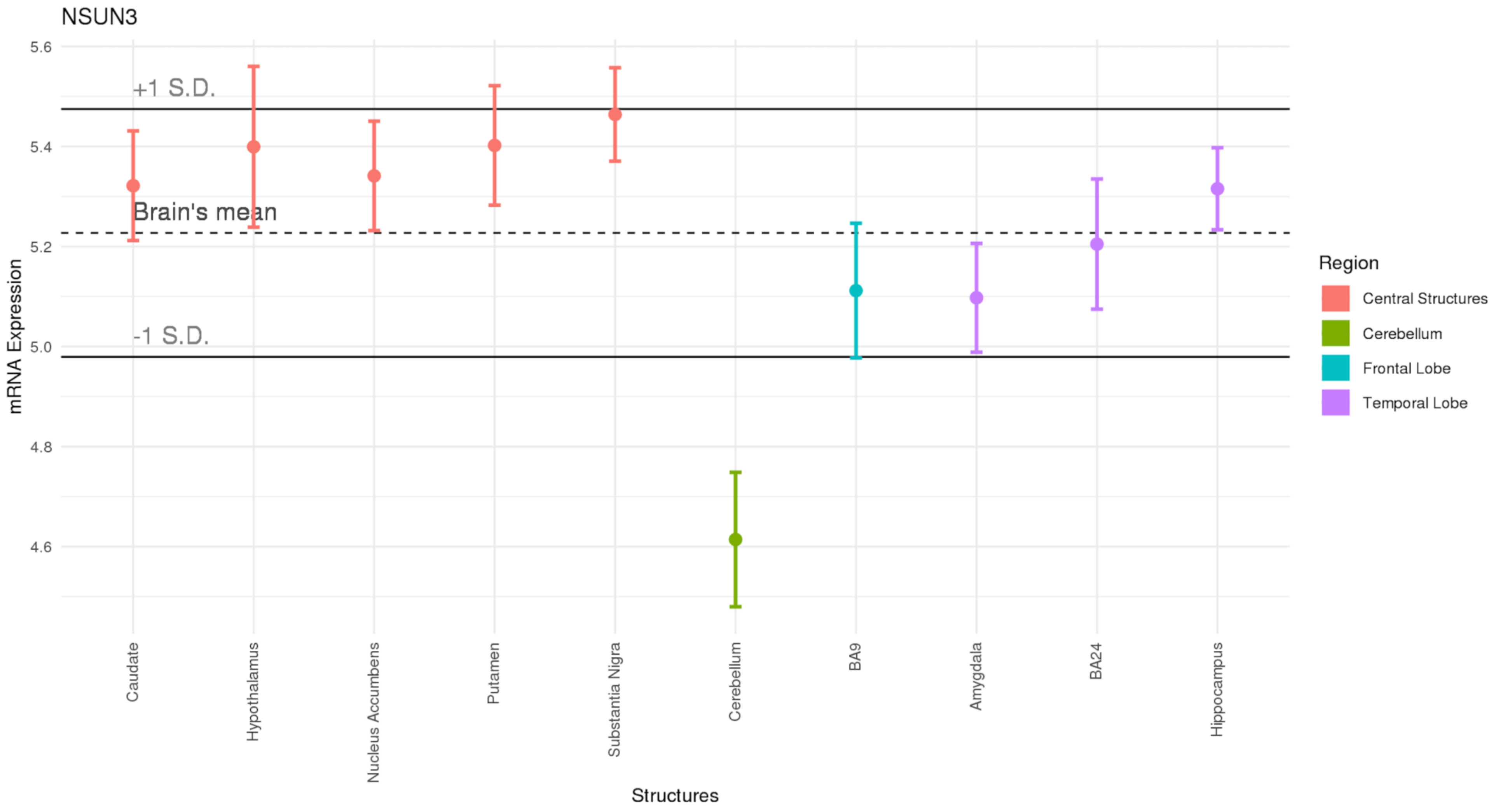
NSUN3
+1 S.D.
Brain's mean
-1 S.D.
mRNA Expression
5.6
5.4
5.2
5.0
4.8
4.6
Caudate
Hypothalamus
Nucleus Accumbens
Putamen
Substantia Nigra
Cerebellum
BA9
Amygdala
BA24
Hippocampus
Structures
Region
Central Structures
Cerebellum
Frontal Lobe
Temporal Lobe

**h**

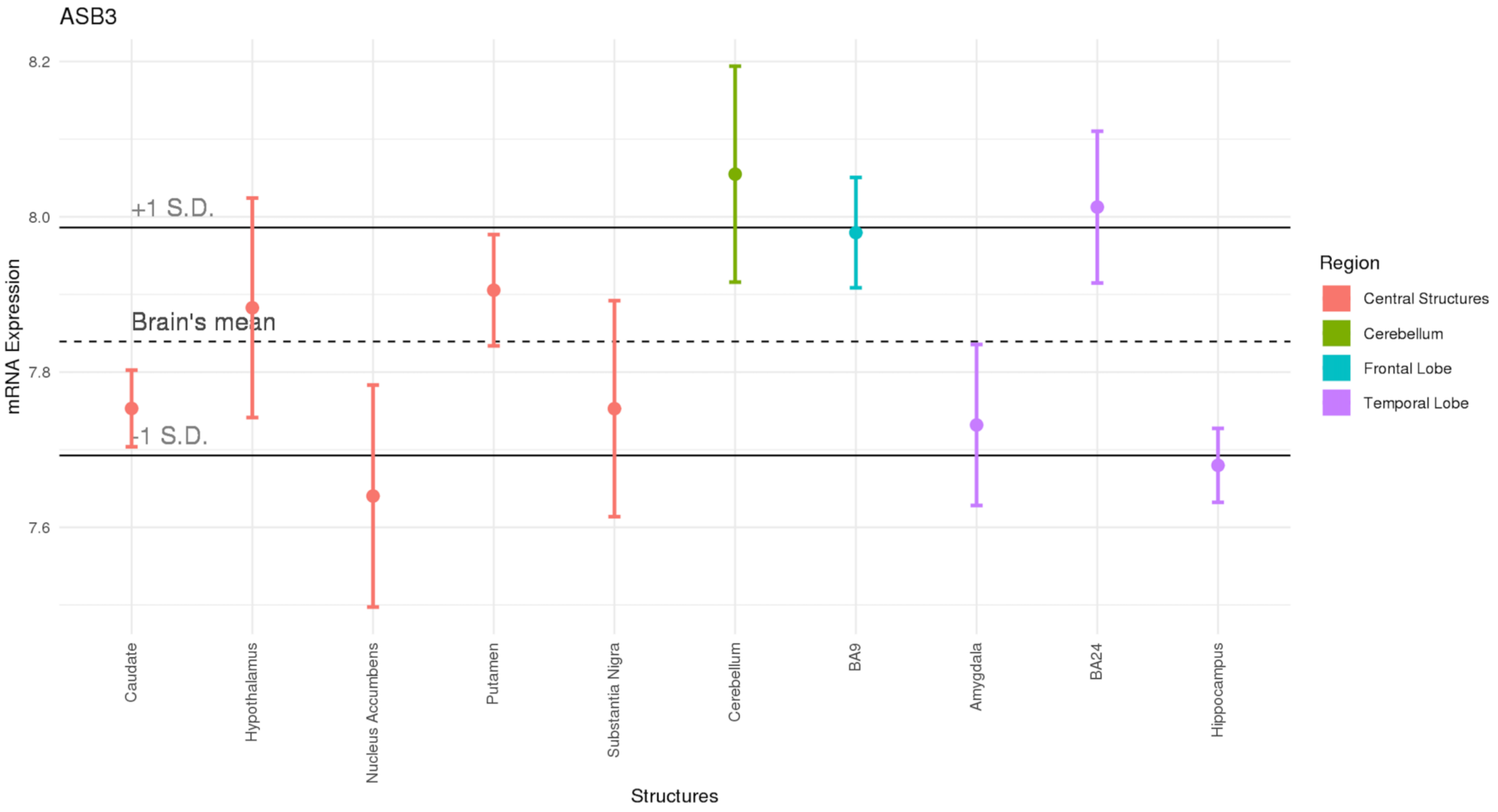
ASB3
8.2
8.0
7.8
7.6
+1 S.D.
Brain's mean
-1 S.D.
mRNA Expression
Caudate
Hypothalamus
Nucleus Accumbens
Putamen
Substantia Nigra
Cerebellum
BA9
Amygdala
BA24
Hippocampus
Structures
Region
Central Structures
Cerebellum
Frontal Lobe
Temporal Lobe

**i**

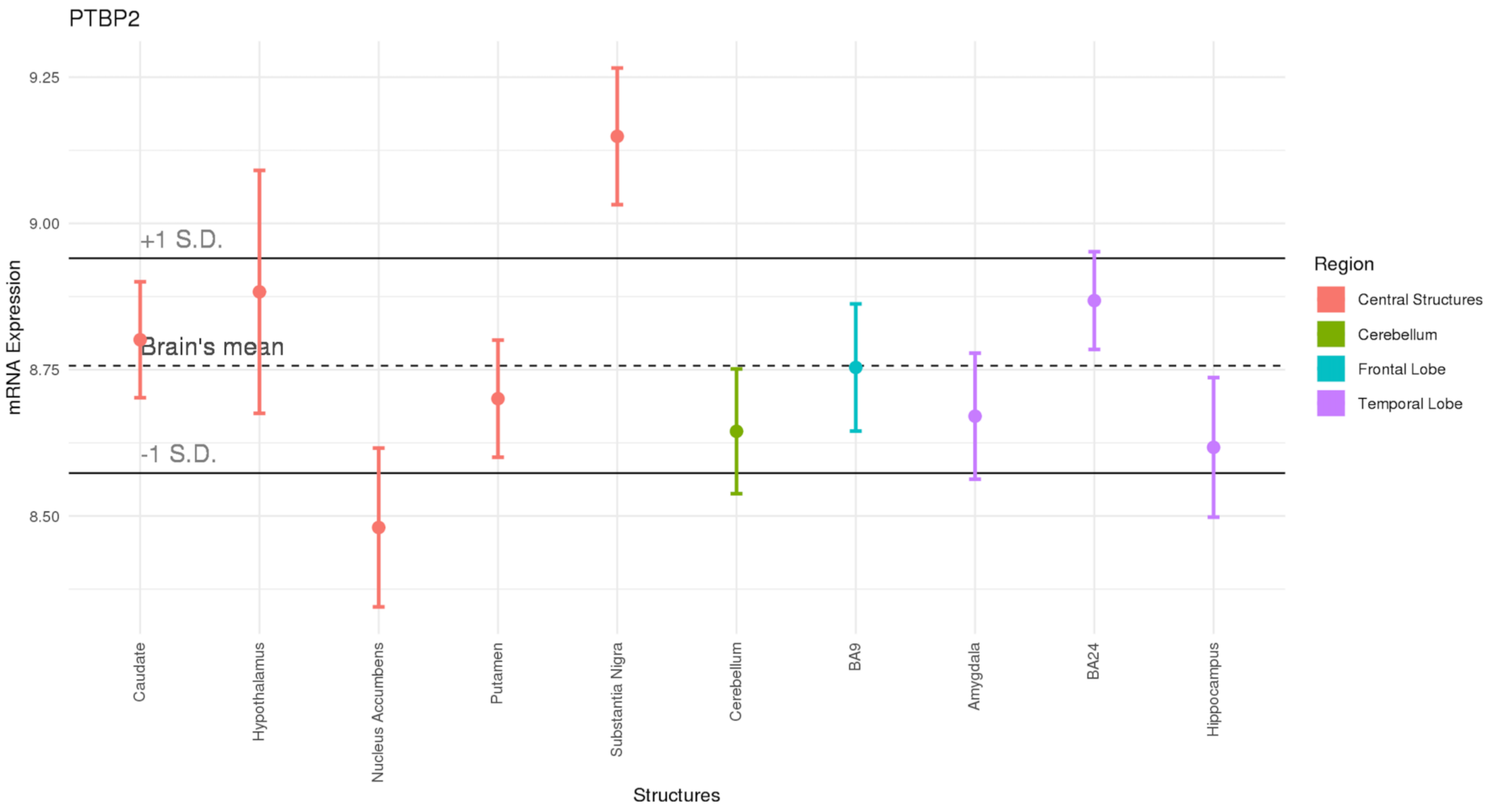
PTBP2
9.25
9.00
8.75
8.50
mRNA Expression
+1 S.D.
Brain's mean
-1 S.D.
Caudate
Hypothalamus
Nucleus Accumbens
Putamen
Substantia Nigra
Cerebellum
BA9
Amygdala
BA24
Hippocampus
Structures
Region
Central Structures
Cerebellum
Frontal Lobe
Temporal Lobe

**Supplementary Figure 5**: **(a)** Heat map of gene expression for each of the 9 AN risk genes across 30 different tissue types from the GTEx dataset, and **(b)** differential expression of the AN gene set across 48 tissue types from the GTEx dataset. -log pa values represent the probability of the hypergeometric test.

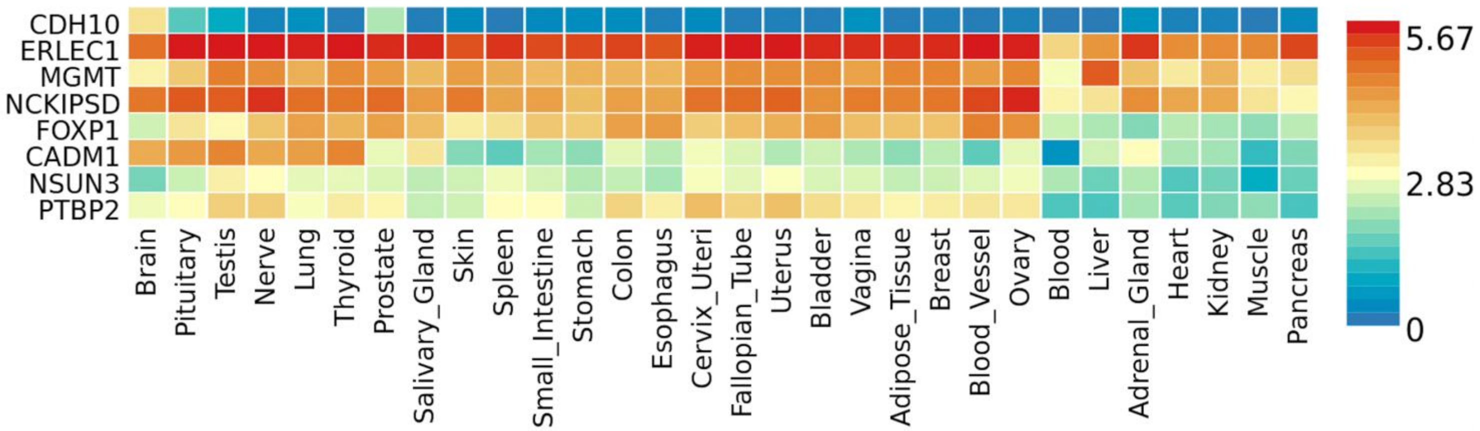


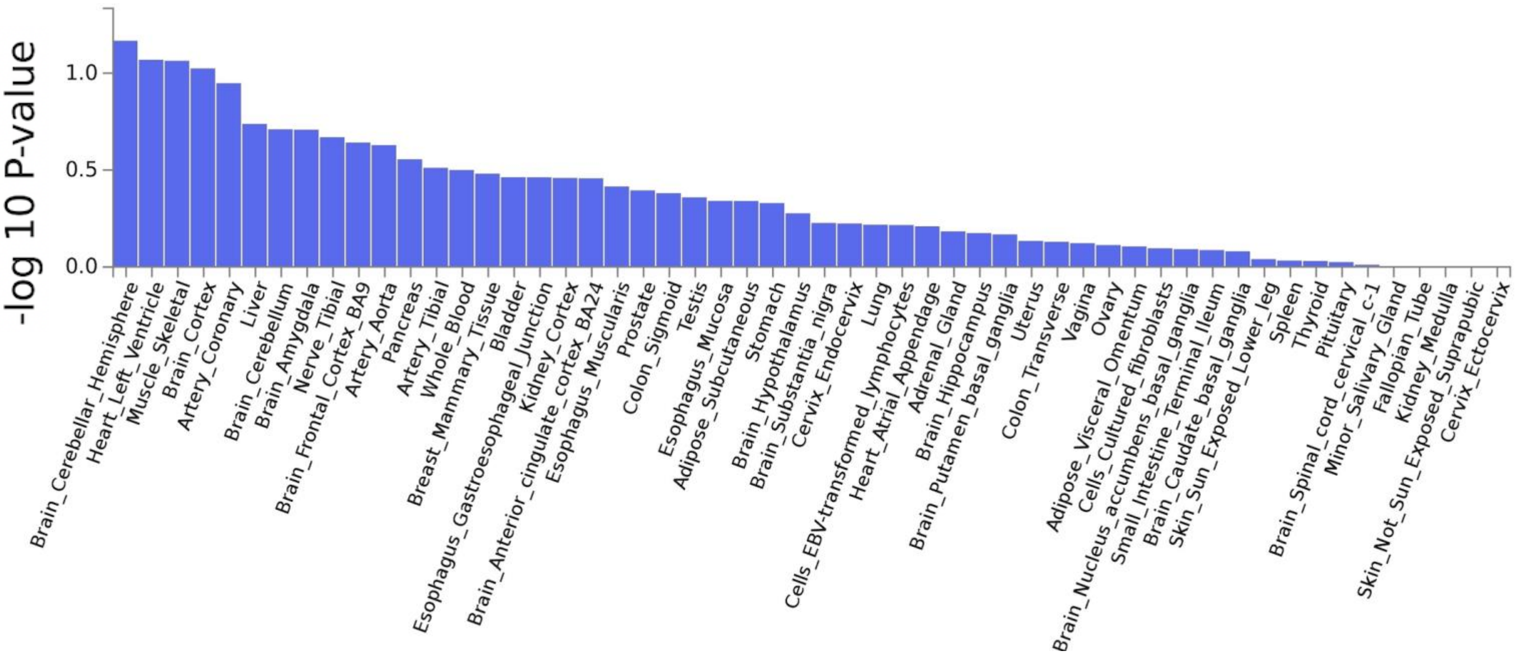
